# Salt warning labels in the out-of-home food sector: online and real-world randomised controlled trials

**DOI:** 10.1101/2025.01.13.25320447

**Authors:** Rebecca Evans, Jane Brealey, Natasha Clarke, Jennifer Falbe, Amy Finlay, Andrew Jones, Paula Thorp, Beth Witham, Rozemarijn Witkam, Eric Robinson

## Abstract

**Background:** High salt intake is a significant cause of diet-related disease. The salt content of much of the food provided in the out-of-home food sector (OOHFS; e.g., restaurants) can be excessive, but policy options to address this are lacking. An emerging policy approach with limited evidence from randomised controlled trials (RCTs) is the use of ‘high in salt’ warning labels on restaurant menus.

**Methods:** UK adults participated in an RCT to test the perceived message effectiveness (PME) and impact on hypothetical food choice of salt warning labels in packaged food and restaurant scenarios (*Study 1*). Next, the same outcomes were examined in a real-world RCT. Customers selected, consumed and purchased meals in a restaurant from menus with vs. without salt warning labels (*Study 2*).

**Findings:** Study 1 (N = 2391) demonstrated that salt warning labels were perceived by consumers as being effective in discouraging high salt intake and reduced hypothetical salt selection vs. control across both packaged food (on average −0.08g [95% CI −0.12 to −0.04] per item) and restaurant (on average − 0.26g [95% CI −0.43 to −0.10] per meal) scenarios. Study 2 (N = 454) replicated findings from Study 1, for perceived effectiveness and reduced salt selection vs. control (−0.54g [95% CI −0.83 to −0.25] per meal) in a real-world restaurant setting.

**Interpretation:** Salt warning labels on restaurant menus are a promising policy option to address excessive salt intake in the OOHFS.

**Funding:** National Institute for Health and Care Research Oxford Health Biomedical Research Centre (NIHR203316); European Research Council (8031940).

## Research in context

### Evidence before this study

We searched studies on the impacts of nutrient warning labels in PubMed from 1^st^ January 2000 to 5^th^ November 2024 using the search terms (“nutrient warning label*” OR “nutrient warning*” OR “chile* food label*” OR “salt warning label*” OR “salt warning*” OR “salt label*” OR “sodium warning label*” OR “sodium warning*” OR “sodium label*”) based on titles and abstracts. We identified 47 articles.

Front-of-pack (FoP) nutrient warning labels are currently implemented mandatorily in several South American countries, including Chile (since 2016), Brazil and Mexico, and further afield in Canada and Israel. Meta-analytic evidence from predominantly hypothetical or laboratory-based experiments indicates that FoP nutrient warning labels are perceived to be effective by consumers and reduce the selection of (i) less healthy products and (ii) energy and nutrients of concern (salt [-0.08g, 95% CI −0.15, −0.02], saturated fat, sugar). Limited research has examined menu-based ‘high in salt’ warning labels in the out-of-home food sector (OOHFS). In a series of online randomised controlled trials (RCTs) with US consumers, salt warning labels increased negative health perceptions of the food, and reduced hypothetical average salt ordered (by 0.05-0.2g). Observational research in the US (New York City) suggests that implementation of salt warning labels in the OOHFS is associated with decreases in the total salt content of meal purchases from full-service restaurants.

### Added value of this study

There are a limited number of RCTs examining the impact of salt warning labels on menus in the OOHFS, and no trials have been conducted in real-world settings. In the UK, although there is government interest in food labelling approaches, there is a lack of research to guide policy development. In the present research, co-design methods were used to develop salt warning labels that could be used in the UK OOHFS, before testing their perceived effectiveness and impacts on food choice in (i) an online RCT and (ii) a real-world RCT in a restaurant.

### Implications of all the available evidence

The present research is the first to demonstrate that salt warning labels on menus reduce the salt content of hypothetical selections and in real-world restaurant settings. Importantly, findings indicate that salt warning labels are effective in a UK context and should be considered by the UK Government as a public health policy in the OOHFS to reduce population-level salt intake.

## Introduction

Excess salt intake is associated with a range of negative health outcomes and is responsible for an estimated 1.89 million deaths worldwide each year (1). In England, the average adult consumes 40% more salt than the recommended 6g per day (2). This overconsumption is driven by pre-prepared food and food eaten in the out-of-home food sector (OOHFS), much of which is high in salt (2,3).

An emerging public health policy which has been adopted in several countries is the mandatory implementation of front-of-pack (FoP) nutrient warning labels. Nutrient warning labels communicate that a food item contains a “high” amount of energy or a specific nutrient of concern (e.g., salt). The first example of FoP warning labels was in Chile, which implemented the labels in 2016 for packaged foods high in calories, saturated fat, sugar, and/or salt alongside complementary nutrition policies (4). Meta-analytic evidence from predominantly hypothetical and laboratory-based experiments suggests that FoP nutrient warning labels are perceived to be effective by consumers, reduce probability of selecting less healthful products, and reduce hypothetical purchase of energy and nutrients of concern, including salt (−0.08g, 95% CI −0.15, −0.02) (5,6). Post-implementation evaluations of Chile’s FoP warning label policy also suggest this approach may be effective in terms of reducing purchase of labelled items, and nutrient of concern content via reformulation (7,8).

However, FoP nutrient warning labels do not address a key driver of excessive salt intake – the OOHFS (2). Recent policy developments in the US have resulted in salt (or ‘sodium’) warning labels becoming mandatory in New York City and Philadelphia for chain restaurant menu items that exceed the daily recommended limit (4). In a series of online hypothetical studies with US consumers, salt warning labels increased negative health perceptions of high-salt foods, and reduced hypothetical average salt ordered (9). Observational research from New York City suggests that implementation of salt warning labels was associated with decreases in the total salt content of meal purchases (- 1.31g, 95% CI −2.32, −0.30), but due to the correlational nature of this single study, causal evidence for this policy approach is limited(10).

The World Health Organisation has recommended menu labelling as a key measure to reduce sodium intake (1). The UK Government has announced intentions to introduce new food labelling policy and has a long-term interest in reducing salt in the UK diet (11–13), but research is needed to guide policy development. To date, there is no research testing salt warning labels in a UK context, and no real-world RCTs on the impact of salt warning labels in the OOHFS. In the present research, we examined the potential effectiveness of salt warning labels on menus in the UK OOHFS. Using co-design methods, we first developed a range of salt warning labels and tested their impact on perceived message effectiveness (PME) and hypothetical food choice in an online randomised controlled trial (RCT) (*Study 1*). Next, we moved to a large real-world RCT in which we examined the impact of salt warning labels on PME and consumer behaviour in a restaurant setting (Study 2).

## Study 1

### Methods

Methods and analysis plans were pre-registered on the open science framework (OSF) (https://osf.io/vyh2x/?view_only=9063c20d2cd14069aeb953eda4ccac27). Since there is a lack of evidence on warning labels in a UK context, in Study 1 we examined salt warning labels in the context of both packaged supermarket food (to replicate limited existing evidence from other countries (5)), and in a restaurant context.

### Participants

Participants were recruited from the online panel provider, Prolific (14). Inclusion criteria were current UK resident, aged 18 years and over, fluent in English, purchase supermarket sandwiches and savoury snacks and eat out at/order from restaurants at least monthly. Exclusion criteria were pregnant/breastfeeding, partaking in fast/restrictive eating at time of participation, and significant dietary restrictions/intolerances (e.g., vegan, vegetarian, gluten-free). We used stratified sampling to ensure the sample was representative of the UK in terms of age, sex, and education (15–17) (see Appendix A for the stratification plan). Participants were paid at a rate of £6/hr.

### Design

The study was an online randomised controlled trial design. Participants were randomised (between-subjects) to one of four salt warning labelling conditions, or a control condition (see Table 1; also see Appendix B for a summary of key conclusions from the Patient and Public Involvement (PPI) sessions regarding label design) and made hypothetical food choices in packaged supermarket food and restaurant scenarios (within-subjects).

**Table 1:**
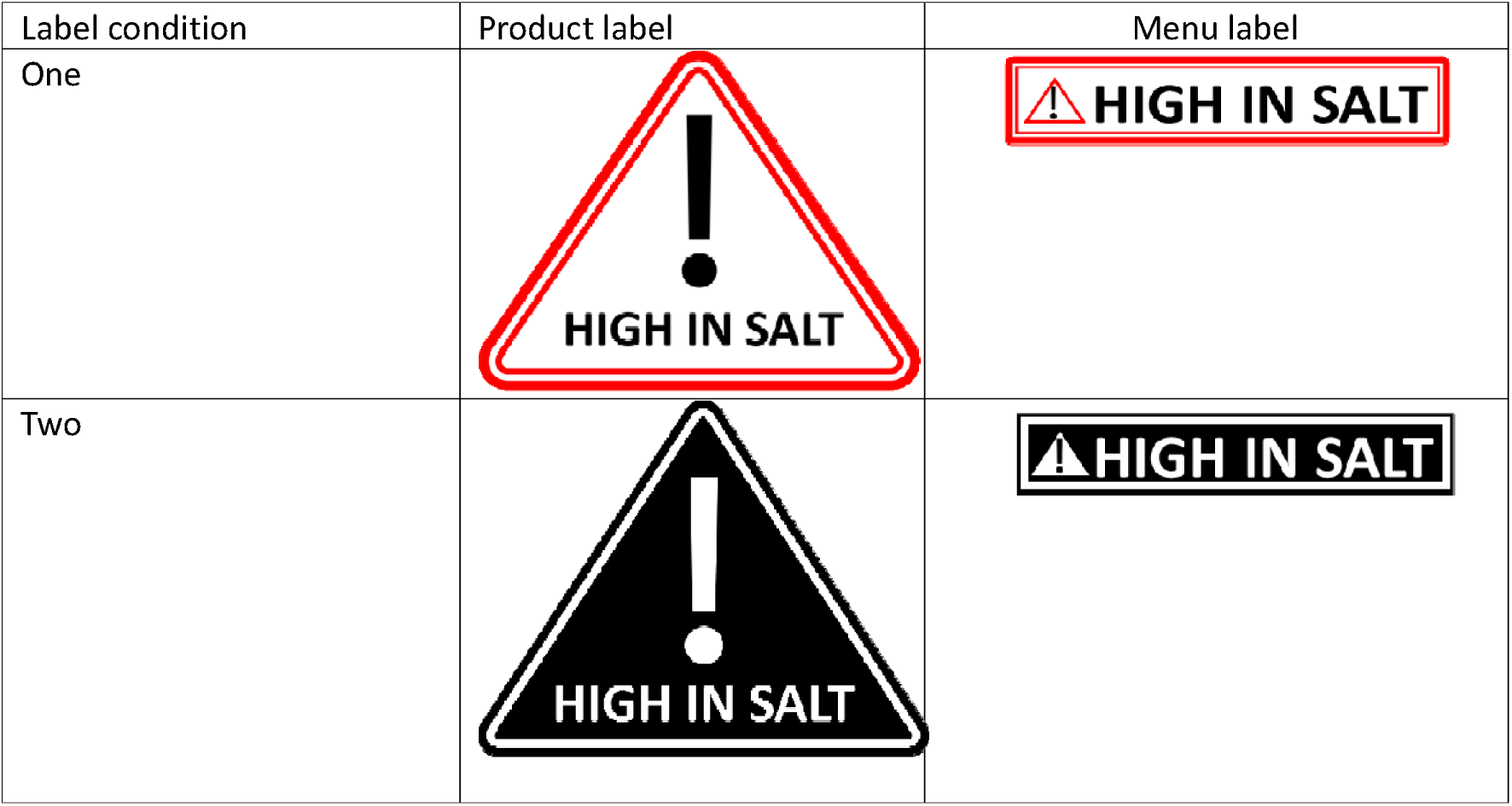
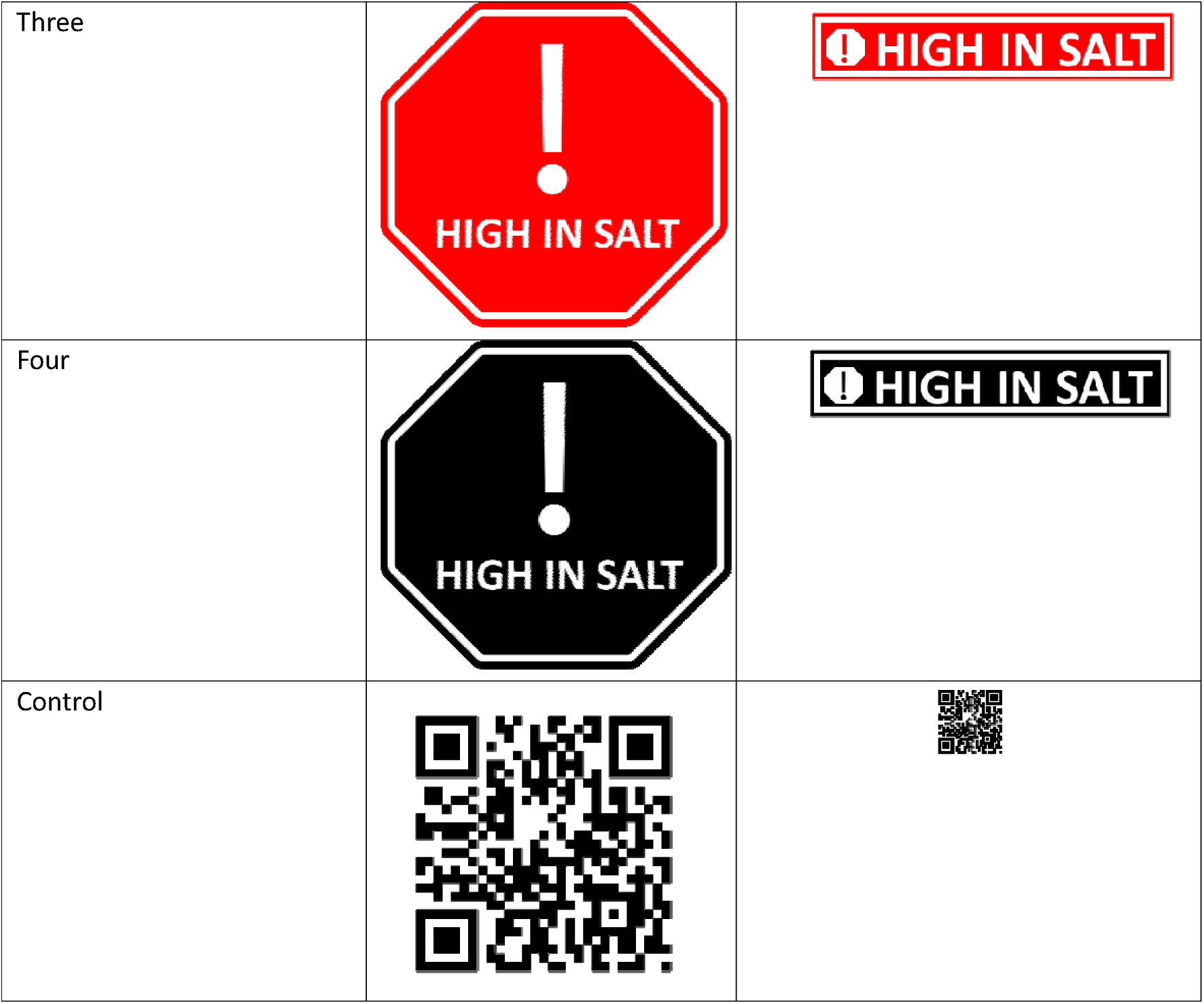
Associated product and menu label conditions.

**Table 2:**
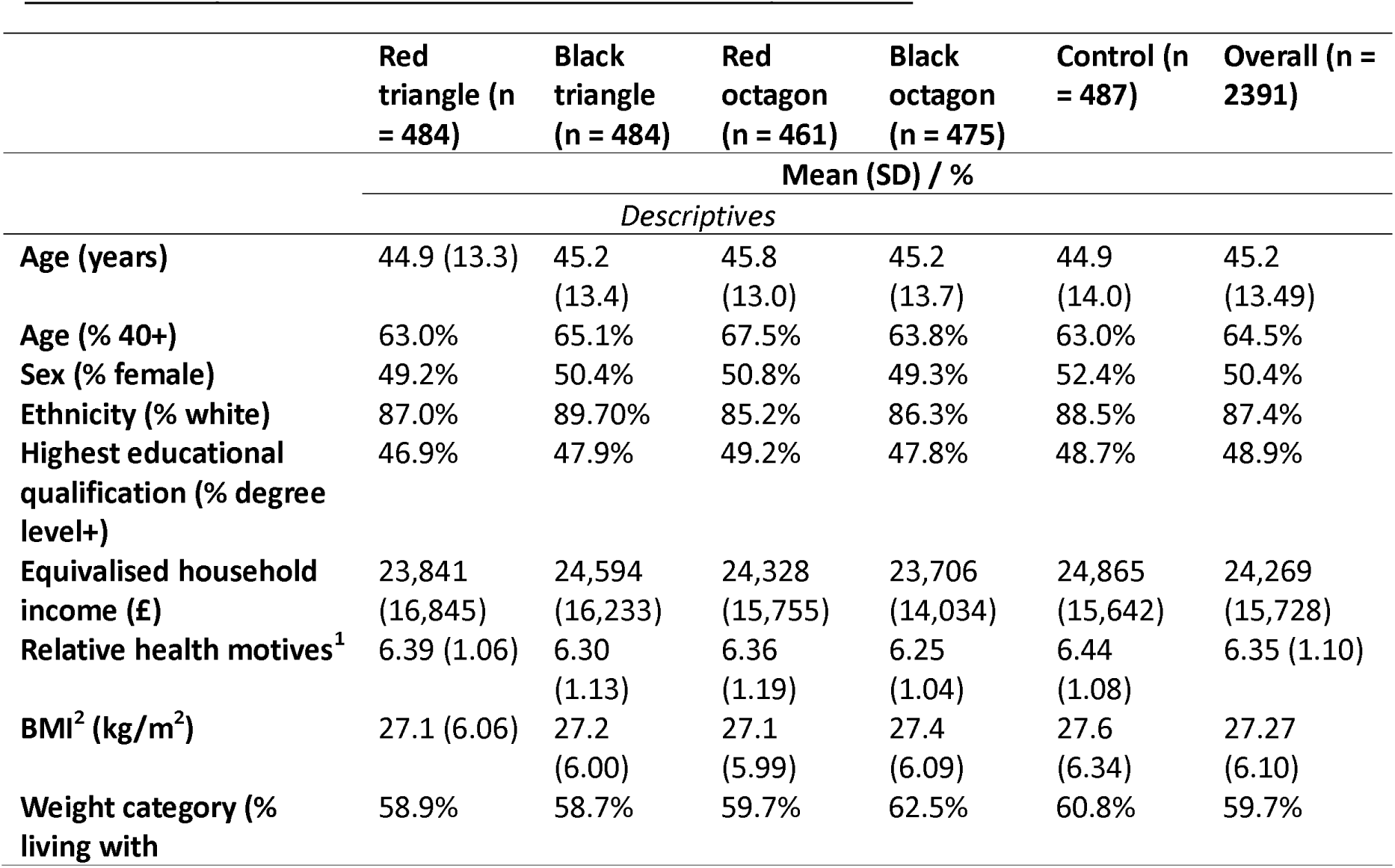
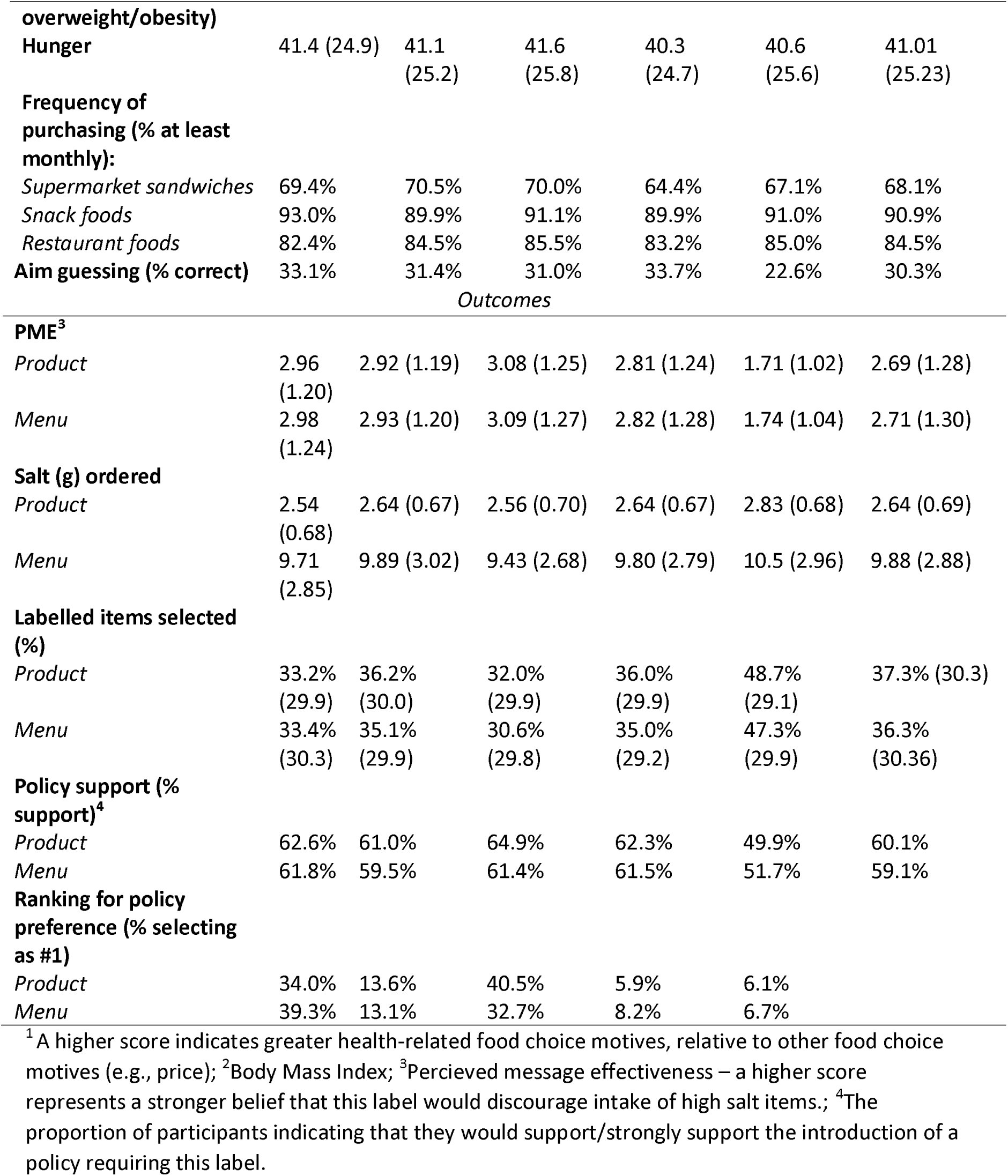
Descriptive statistics and outcomes overall and by condition.

**Table 3:** Descriptive statistics and outcomes overall and by condition.

|  | Labelling condition (n = 238) | Control condition (n = 216) | Overall (n = 454) |
| --- | --- | --- | --- |
|  | Mean (SD) / % |  |  |
|  | *Descriptives* |  |  |
| **Age (y)** | 50.98 (15.68) | 48.44 (16.44) | 49.77 (16.08) |
| **Sex (% male)** | 40.4% | 50.5% | 45.2% |
| **Highest educational qualification (% degree level+)** | 50.2% | 54.7% | 52.3% |
| **Employment status (% full-time)** | 40.0% | 45.8% | 42.8% |
| **Ethnicity (% white)** | 90.6% | 89.7% | 90.2% |
| **BMI (kg/m<sup>2</sup>)<sup>1</sup>** | 26.67 (4.28) | 27.48 (6.12) | 27.06 (5.26) |
| **Weight category (% living with overweight or obesity)** | 60.6% | 60.5% | 60.5% |
| **Frequency of eating out (% at least monthly)** | 91.5% | 90.2% | 90.9% |
| **Aim guess (% yes)** | 20.0% | 0.0% | 10.5% |
| **Health motives<sup>2</sup>** | 2.78 (0.68) | 2.74 (0.64) | 2.76 (0.66) |
| *Outcomes* |  |  |  |
| **PME<sup>3</sup>** | 2.32 (1.16) | 1.32 (0.66) | 1.85 (1.08) |
| **Labelled item ordered (% yes)** | 45.0% | 52.3% | 48.5% |
| **Total salt ordered (g)** | 3.77 (1.52) | 4.31 (1.63) | 4.03 (1.60) |
| **Salt intake (g)** | 3.49 (1.45) | 3.95 (1.56) | 3.71 (1.52) |
| **Salt awareness (% yes)<sup>4</sup>** | 48.9% | 7.0% | 29.0% |
| **Policy support (% support)<sup>5</sup>** | 64.3% | 67.8% | 65.9% |

<sup>1</sup>Body Mass Index, 54 participants did not provide height and/or weight data; <sup>2</sup>A higher score indicates greater health-related food choice motives; <sup>3</sup>Perceived message effectiveness – a higher score represents a stronger belief that this label would discourage intake of high salt items; <sup>4</sup>The proportion of participants indicating that they thought about the salt content of menu items before making their selection; <sup>5</sup>The proportion of participants indicating that they would support/strongly support the introduction of a policy requiring this label

### Food selection tasks

Participants were instructed to make food selections in packaged supermarket food and restaurant scenarios. In the packaged food scenarios, participants were asked to select a sandwich, pack of crisps and a savoury snack. In the restaurant scenarios, participants were asked to choose a main course for an evening meal from three restaurant menus (based on popular UK restaurants of different cuisine types – Italian, Japanese, and fried chicken). In each scenario, participants chose one item from six options, half of which were high in salt. Products were classed as high in salt if they contained > 1.5g of salt per 100g or >1.8g per portion (as per the voluntary red traffic light threshold in the UK for packaged food; (18,19)). Restaurant meals were categorised as high in salt if they exceeded 50% of the recommended daily limit for salt intake in the UK (> 3g) (20). We used a 50% threshold rather than the 100% threshold implemented in some US cities because the 100% threshold would only cover a minority of high salt items in UK chain restaurants (approximately 4% of all menu items), as salt content is typically lower than in US restaurants (3).A 50% threshold is also closer to current UK traffic light thresholds for high salt. Label reference information was included at the foot of each selection page, e.g. “[Label] indicates that the [product/meal] is high in salt…”. See Appendix C for included food items.

### Measures

#### Demographics and baseline measures

Participants were asked to report their sex, age and ethnicity. Self-reported height and weight data were collected to calculate body mass index (BMI) and weight category (21). Two measures of socioeconomic position (SEP) were collected, (i) highest education level, and (ii) equivalised household income (i.e., adjusted to account for differences in household size and composition) (22).

Baseline levels of hunger were measured using visual analogue scales ranging from 1 – 100 (“Not at all” to “Extremely”). Frequency of (i) supermarket sandwich and snack purchase, and (ii) eating at/ordering from restaurants were measured on a 5-point Likert scale (“Not in the last year” to “Three times per week or more”).

### Relative health motives

The Food Choice Questionnaire (23) was administered to measure factors that may influence participants’ food choices. We calculated the relative importance of the health motive by dividing the participant’s absolute health motive score by their mean score on all items (24).

### Perceived message effectiveness (PME)

PME was measured using an adapted version of the University of North Carolina (UNC) PME scale (25). Participants answered three questions using a Likert scale ranging from 1 – 5 (“Not at all” to “A great deal”): “This label… (i) makes me concerned about the health effects of consuming items high in salt, (ii) makes consuming items high in salt seem unpleasant, (iii) discourages me from wanting to consume items high in salt”. The mean response to the three items was calculated. The scale has been validated and is predictive of real-world behaviour (25–30).

### Food choices

Food choices were measured as (i) whether a labelled item was selected (No = 0, Yes = 1) converted to a percentage and (ii) the total salt in food selected (g). This was calculated across (i) the three packaged food scenarios and (ii) the three restaurant scenarios.

### Procedure

Participants completed demographic information and baseline measures. Participants were then randomly assigned to one of the labelling conditions and completed the packaged supermarket food and restaurant ordering scenarios (counter-balanced).

Following this, participants were shown an image of (i) a packaged product and (ii) a menu item featuring their assigned label (counter-balanced). They were asked about awareness, perceived effectiveness, knowledge gain, and influence of the labels, and support for mandating them. Finally, participants provided more detailed demographic information and answered a question regarding study aims. Two attention checks and one consistency check were included. See Appendix D for detailed study flow and Appendix E for additional measures.

### Analysis

#### Power

It was calculated that a minimum sample of 2180 participants (436 per condition) would provide sufficient power (80%) to detect small between-condition effects (d = 0.19) of warning labels on PME, salt (g) selected, and % of labelled items selected (5,9). We aimed to recruit up to 2500 participants (500 per condition) to account for potential loss of up to approximately 15% due to data exclusions (see Appendix E for power analysis full detail and effect size justification).

#### Primary

A mixed ANOVA was conducted to examine the effect of label condition and selection scenario on ratings of PME. Label condition was entered as the between-subjects factor, and scenario (packaged food, menu) was entered as the within-subjects factor. Age (18 – 39, 40+), sex (male, female), and education (no University degree, degree level or above) were included as covariates to account for stratification. Relative health motives were entered as a covariate as we assumed motives would be predictive of outcomes (e.g., health motivated individuals may be more likely to believe a menu with labels will change their behaviour) (31).

#### Secondary

Mixed ANOVAs were conducted to examine the effect of label condition and selection scenario on (i) the proportion of labelled items selected (%) and (ii) salt (g) ordered. The same factor structure as above was used. Values for salt ordered were standardised (converted to z scores) within each scenario, due to differences in the range of the salt content of items in the product and menu scenarios.

#### Sensitivity

Primary and secondary analyses were reconducted after removing participants who correctly guessed study aims.

## Results

After removal of participants for questionnaire incompleteness (N = 2), failing an attention check (N = 18), consistency check (N = 54), or implausible BMI values (<10 or >60kg/m^2^, N = 84), the final analytic sample was N = 2391. See Appendix D for participant flow.

### Primary analysis

#### PME

There was a significant main effect of label condition on PME (F (4, 4758) = 224.31, p < .001, partial eta^2^ = 0.16, 95% CI [0.14, 1.00]). There was no significant main effect of scenario, and no condition*scenario interaction. Post-hoc analysis indicated that all salt warning labels were rated as having significantly greater perceived effectiveness for changing behaviour than the control condition (all p<.001). The red octagon label was also rated as significantly more effective than the black triangle (p = 0.03), and the black octagon (p <.001). See Figure 1.

**Figure 1:**
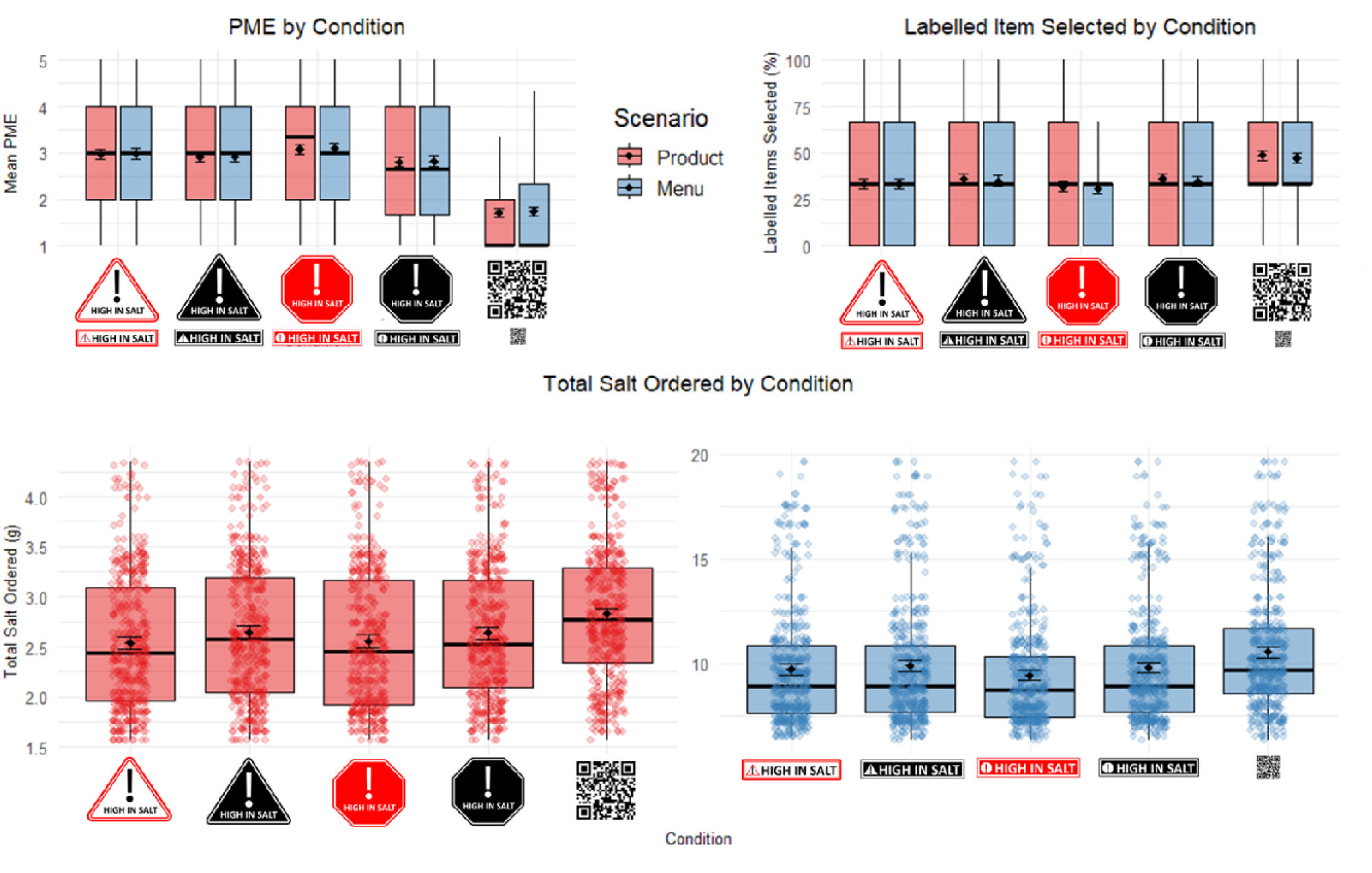
Boxplots to show PME, salt ordered (g), and the proportion of labelled items selected (%) by condition

### Secondary analysis

#### Salt (g) ordered

There was a significant main effect of label condition on salt ordered (F (4, 4758) = 23.47, p <.001, partial eta^2^ = 0.02, 95% CI [0.01, 1.00]). There was no significant main effect of scenario, and no condition*scenario interaction.

Post-hoc analysis indicated that all salt warning labels resulted in significantly less salt ordered compared to the control condition (all ps <.001). This reduction was on average −0.26g (95% CI −0.43 to −0.10) per restaurant meal, and −0.08g (95% CI −0.12 to −0.04) per packaged supermarket item, relative to the control, based on raw mean differences. The red octagon also resulted in significantly less salt ordered relative to the black triangle (p = .02). See Figure 1.

#### Labelled items selected (%)

There was a significant main effect of label condition on the proportion of labelled products that were selected (F(4, 4758) = 49.56, p <.001, partial eta^2^ = 0.04, 95% CI [0.03, 1.00]). There was no significant main effect of scenario, and no condition*scenario interaction.

Post-hoc analysis showed that all salt warning labels significantly reduced the proportion of labelled items selected relative to the control condition (all p <.001). The red octagon label also resulted in significantly fewer labelled items selected compared to the black triangle label (p = .02). See Figure 1.

Graph displays unstandardized salt values to aid interpretation. Boxes represent the interquartile range (IQR). The line inside the box is the median, and the dot inside the box is the mean with error bars representing 95% confidence intervals. Whiskers represent the range of most of the data (within 1.5 times the IQR). Coloured dots represent individual data points.

See Appendix E for full results, including sensitivity and exploratory analyses. We observed no deviation in findings from primary and secondary analyses when aim guessers were removed, or when study aim awareness was included as a covariate. Briefly, there was a significant interaction between condition and health motives, and condition and age in terms of PME. Greater health motives were associated with higher PME ratings across all labels, but this was particularly pronounced for the red labels relative to the black labels. Over 40s also rated the red triangle as significantly more effective than under 40s. Otherwise, there were no other significant interactions between condition and any covariates for any of the primary and secondary outcomes. Awareness, perceived influence, knowledge gain, and policy support was greater for salt warning labels relative to the control, and the red triangle and red octagon labels were favoured by participants.

##### Interim discussion

Findings suggest that salt warning labels are perceived to be effective by UK consumers, are supported as a public health policy, and reduce the proportion of labelled items selected and total salt ordered (hypothetical) for both packaged food and importantly, restaurant food. Study 2 builds on Study 1 by testing the real-world effectiveness of one of the best performing and most highly ranked label in participant preference ratings (“Red triangle”).

## Study 2

Methods and data analysis plans were pre-registered on the OSF (https://osf.io/whzc8/?view_only=e97eb9ff6812457081d6d693e27bf1bd) and ClinicalTrials.gov (ID: NCT06458270).

### Participants

Participants were recruited via targeted social media adverts (Facebook, Instagram) in the local area (Liverpool, UK), and the study was described as being about dining habits in restaurants. Interested participants were asked to complete an online screening questionnaire. Inclusion and exclusion criteria were less restrictive than the previous study (vegetarians and those with dietary requirements that did not substantially constrain choice were permitted), due to the real-world nature of the study. Participants registering interest online could bring a maximum of nine co-diners (who would also participate) to the restaurant. We aimed to recruit a sample that was representative of the UK in terms of education primarily, and also age and sex (15–17). Participants each received a £25 reimbursement for their time.

### Design

The study used a between-subjects RCT design. The between-subject factor was the menu labelling condition (labelled menu, standard menu). The study was conducted in a full-service independent pub restaurant in Liverpool City Centre which serves a variety of food (e.g., burgers, pies, salads, flatbreads/sandwiches).

## Materials

### Study menus

In the control condition, participants were given the restaurant’s standard menu with no labels. In the experimental condition, participants were given a menu with salt warning labels next to items that were high in salt. All menu items were weighed and sent for nutritional analysis to determine salt content/100g (see Appendix F for menu item nutritional information). Labels were placed below the name of menu items that exceeded 50% of the adult daily recommended limit for salt in the UK (>3g, 5/16 items) (20,32). Labels were the same height as the menu item text (33), and reference text was included at the foot of the menu “[Label] indicates that the salt content of this item is higher than 50% of the daily recommended limit (6g per day). High salt intake can increase blood pressure and risk of heart disease and stroke”. Drink options (including alcohol), were presented on a separate menu. See Appendix G for study menus.

### Measures

The measures used in Study 2 were largely the same as in Study 1. Any deviations are described below.

Employment status was collected in place of equivalised household income to reduce questionnaire length.

### PME (25)

Phrasing was adapted so participants were asked to consider the statements in relation to the menu ordered from, e.g. “The menu made me concerned about the health effects of consuming items high in salt”.

### Health motives (23)

Mean health motives score was computed, rather than relative health motives. This was because only the health motives questions (6 items) were displayed to participants to reduce questionnaire length.

### Food choices

Ordering of a labelled item was measured as a categorical variable (Yes = 1, No = 0) as there was only one ordering scenario. There was no limit on the number of items each participant could order.

In addition to salt selection, salt intake was determined by trained nutritionists based on (i) the salt content of the order and an estimation of the proportion of the meal that was consumed (from pre- and post-intake photos), and (ii) participant self-report of meal sharing and addition of condiments (as a percentage/teaspoon estimate). A random 10% of photos were second-coded to ensure consistency. See Appendix F for details on the procedure for calculating salt intake.

### Procedure

Participants attended one lunch session at the restaurant from Monday – Saturday between 12 and 3pm. All participants were asked to verbally consent to taking part in the study, and were given the opportunity to ask questions before the study began. Participants attending together were sat at the same table and randomised to the same menu condition.

Participants were given their assigned menu and asked to write their order on a form. The researchers received training from the restaurant and acted as waiting staff; they took orders and delivered/collected meals alongside taking covert pre- and post-intake photos. After finishing their meal, participants were asked to fill out a survey on an iPad to measure study aims, meal sharing/condiments added, demographics, salt awareness, perceived message effectiveness, and policy support. Participants then paid for their meal. The next morning participants received an email prompting them to record all food consumed after the restaurant visit (up to midnight the same day) using a validated dietary recall assessment tool (Intake24). See Appendix H for detailed study flow. Participants were reimbursed for their participation 2-3 weeks after taking part (on average).

### Analysis

#### Power

It was calculated that a minimum sample of 260 participants (130 per condition) would provide sufficient power (80%, two-tailed) to detect small-medium effects (d = 0.35) of the salt warning label on PME, salt ordered, and salt intake using linear models (5,9). Effect size estimates have a degree of uncertainty, particularly when no previous real-world research has been conducted testing salt warning labels in a restaurant. To balance the trade-off between costs (e.g., in research time) and likelihood of the study providing convincing evidence, we conducted planned interim primary and secondary analyses once we had 130 completers in each condition (34). At the interim analysis, we found clear evidence of between-condition effects on PME, and evidence of directional effects on salt ordered, as anticipated. For salt ordered, it was estimated that approximately 450 participants (225 per condition) would be required to observe a detectable effect at p <.025 (p value halved due to multiple testing (34)). Therefore, as specified in our pre-registered protocol we continued recruitment, with the aim of achieving a minimum sample size of 450 by the end of the study period.

#### Primary

A linear regression was used to examine the effect of menu condition on PME. Age, sex, SEP (degree/no degree) and health motives were entered as covariates, based on Study 1 findings (see Appendix E) (31). There was no evidence of table clustering in terms of PME (Λ = 3.19, p = .07, Intraclass Correlation Coefficient [ICC] = 0.04), therefore table group was not included as a random effect in the final model.

#### Secondary

Linear mixed models were used to examine the effect of menu condition on (i) salt ordered and (ii) salt intake. The same covariates as above were included. There was evidence of clustering in terms of salt ordered (Λ = 9.94, p = .002, ICC = 0.11), and salt consumed (Λ = 8.53, p = .004, ICC = 0.10) and therefore table group was included as a random effect in both models.

A binomial logistic regression was used to examine the exposure of salt labelling (vs not) on the odds of ordering a labelled item. The same covariates as above were included. There was no evidence of table clustering in terms of ordering a labelled item (Λ = 2.98, p = .08, ICC = 0.10), therefore table group was not included as a random effect in the final model.

#### Sensitivity

Primary and secondary analyses were reconducted after removing participants who correctly guessed study aims. We also reconducted a secondary analysis by including salt purchased in place of salt ordered as the outcome variable, to check for differences (as in a small number of instances ordered food was unavailable).

## Results

Full data were removed for participants who did not meet study inclusion criteria (N = 2) or were exposed to the wrong menu condition in error (N = 9), resulting in a final analytic sample of N = 454. Order and intake data were excluded for participants who were told that specific menu items were not available in advance of them making their order (N = 2). Post-meal questionnaire data were excluded for participants who failed the attention check (N = 5), and data regarding label influence specifically were excluded for participants who were given a version of the post-meal questionnaire that did not correspond to their condition due to researcher error (N = 7). See Appendix H for participant flow.

For primary and secondary analyses, the threshold of significance was set at p <.025 (p value of <.05 halved due to interim analyses).

### Primary analyses

#### PME

The overall model was significant and explained approximately 27% of variance in PME (adjusted R^2^ = 0.27, F [5, 443] = 34.34, p <.001). The labelled menu was rated to have significantly higher PME than the standard menu (B = 0.98, SE = 0.09, t = −2.54, p <.001, 95% CI [0.80, 1.15]). See Figure 2.

**Figure 2:**
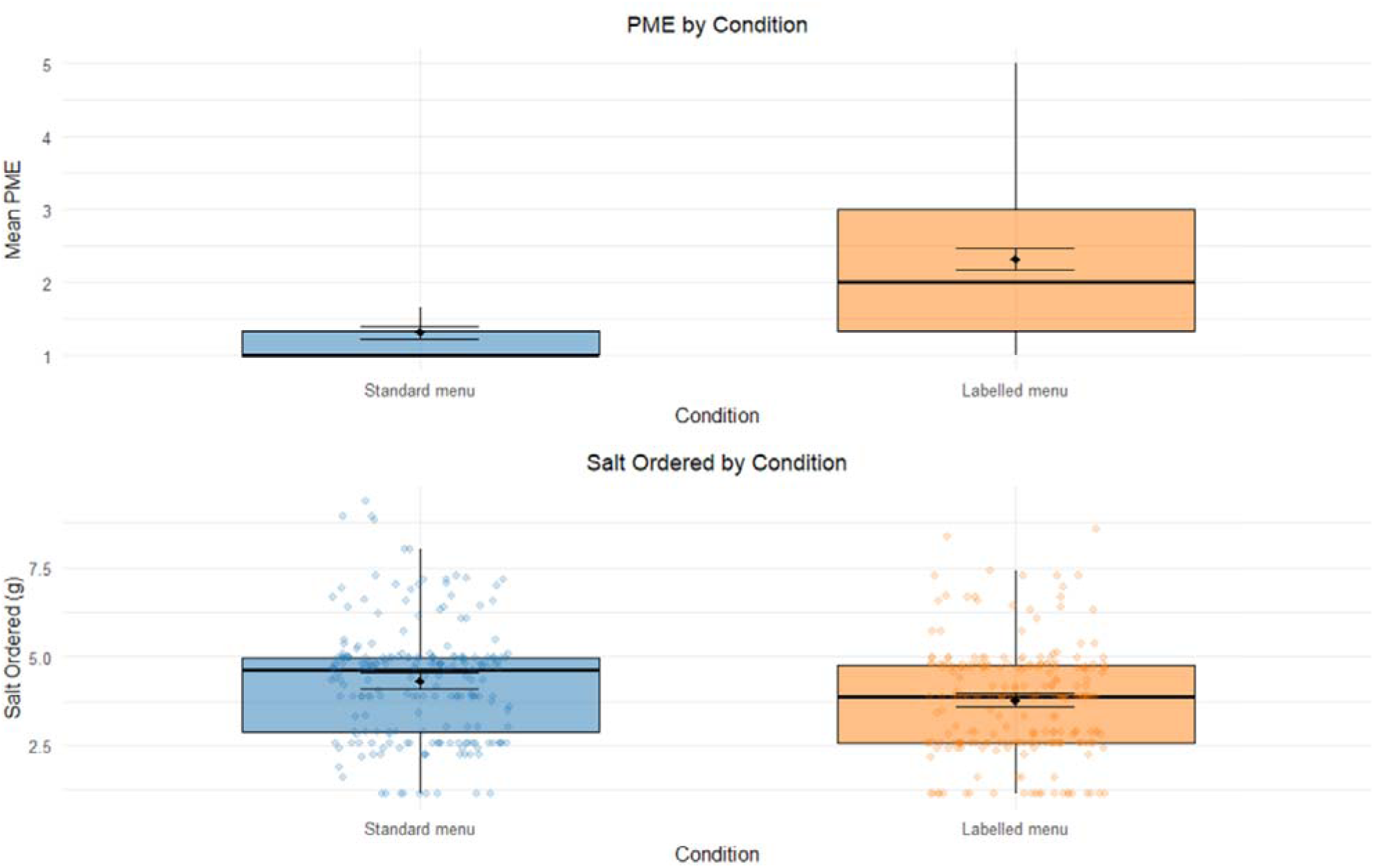
Boxplots to show mean perceived message effectiveness (PME) and salt ordered (g) by menu condition

### Secondary analyses

#### Salt ordered

Significantly less salt was ordered in the labelled menu condition, relative to the standard menu (B = −0.43, SE = 0.17, t = −2.47, p = .01, 95% CI [−0.77, −0.09]). Participants in the labelled menu condition ordered 0.54g (95% CI −0.83 to −0.25) less salt relative to the control based on the raw mean difference. See Figure 2.

#### Salt consumed

There was no significant effect of menu condition on salt consumed (B = −0.31, SE = 0.17, t = −1.87, p = .06 95% CI [−0.63, 0.01]), although, consistent with salt ordered, participants in the labelled menu condition consumed 0.46g (95% CI −0.74 to −0.18) less salt relative to the control based on the raw mean difference.

#### Labelled item ordered

The overall model was significant, explaining approximately 2% of variance in labelled item choice X^2^(5) = 13.65, p =.02, R^2^(McFadden) = 0.02. There was no significant association between menu condition and odds of choosing a labelled item (OR = 0.80, 95% CI [0.55, 1.17], z = −1.13, p =0.26), although, consistent with salt ordered, participants in the labelled menu condition were 20% less likely to order a labelled item relative to the control.

See Appendix I for full results, including exploratory and sensitivity analyses. Removing aim guessers did not substantially impact conclusions, nor did analysis of salt purchase in place of original salt ordered. Briefly, there was no significant impact of condition on later salt intake or other macronutrient intake over the remainder of the day of the study, suggesting no evidence that changes to behaviour in the restaurant were later compensated for. There were no interaction effects between condition and covariates for PME or salt selected. Participants in the labelling condition were significantly more likely to have thought about the salt content of their meal before ordering, relative to the control condition (p <.001).

Boxes represent the interquartile range (IQR). The line inside the box is the median, and the dot inside the box is the mean with error bars representing 95% confidence intervals. Whiskers represent the range of most of the data (within 1.5 times the IQR). Coloured dots represent individual data points.

## Discussion

Two RCTs indicated that salt warning labels on restaurant menus were perceived to be effective by UK consumers and reduced salt ordered in both hypothetical online and real-world settings. Findings on food choice were not moderated by participant age, sex, SEP, or food-choice-based health motives, suggesting that salt warning labels may have equitable effects on consumer behaviour and diet.

Although there is growing policy interest in the use of salt warning labels, research to date has been limited to the US and evidence available comes predominantly from hypothetical choice experiments. The present research is the first to examine the impact of menu-based salt warning labels outside of the US and the first real-world RCT to gauge causal evidence for effectiveness. In a previous online RCT with US consumers making hypothetical food choices, salt warning labels decreased hypothetical salt ordered in a full-service restaurant meal by 0.15g (5,9). Notably, 0.15g is considerably less than in the present research, in which a 0.54g reduction in salt ordered was observed in the real-world experiment. This reduction amounts to approximately ∼10% of the recommended daily limit for salt intake in the UK (6g) (20), suggesting that the introduction of salt warning labels in UK restaurants has the potential to yield substantial public health benefits. Participants in the present studies were largely supportive (66%) of introducing salt warning labels as a policy in the OOHFS and perceived warning labels to be effective at discouraging salt intake. Participants in Study 2 who were exposed to salt warning labels on menus were more likely to report considering the salt content of menu items when ordering.

While an important strength of Study 2 was its real-world setting, there are some limitations to be acknowledged. First, researchers were present and this may have influenced customers to become aware of the study aims and change their behaviour (i.e., demand characteristics). However, analyses in which we removed any participants who guessed study aims produced comparable results to main analyses. Whilst salt ordered was significantly lower in the label condition (p =.01), salt intake was not statistically significant (p= .06). We assume this is due to random variation in food waste and/or minor random error in nutritionist estimates of food waste vs. intake, given that effect sizes for both outcomes were similar (∼0.5g reduction). A further limitation of Study 2 was that we examined effects of salt warning labels in a single restaurant, although it is important to note that in Study 1 we examined hypothetical purchasing behaviour across three different types of restaurant cuisine.

We also cannot generalise our findings to quick-service restaurants; evidence post-implementation of salt warning labels in New York City suggests that reductions in salt ordering were only apparent in full-service restaurants, not quick-service (10). Study 2 can inform further larger-scale RCTs in a variety of real-world restaurant settings, including testing in quick-service settings. Research examining the impact of other nutrient warning labels in the OOHFS may now be valuable, as restaurants also have a substantial number of items that exceed calorie, saturated fat, and sugar recommendations (3). Such research would help to determine whether a more cohesive nutrient warning labelling policy in the OOHFS would be beneficial.

Importantly, this research contributes evidence that can guide policy implementation in the UK (12). If current findings are replicated in larger scale real-world RCTs, the introduction of salt warning labels would be a promising option for the UK Government to reduce population-level salt selection, and tentatively salt intake and associated non-communicable disease. Notably, observed impacts in the present research were via consumer behaviour change alone. If salt warning labelling was introduced as a policy, we presume it is likely that industry reformulation (i.e., reducing salt content of menu items) would result in additional reductions in intake (35).

## Conclusion

Salt warning labels are perceived to be effective by UK consumers and reduced salt selection in both hypothetical online and real-world restaurant scenarios. Salt warning labelling in the OOHFS may be a promising policy option to reduce diet-related disease.

## Data Availability

All data produced in the present study are available upon reasonable request to the authors

