## Appendix for "Salt warning labels in the out-of-home food sector: online and real-world randomised controlled trials"

### **Appendix A – Study 1 sampling plan**

Table A1: Sampling stratification plan

| Variable | Stratification |
| --- | --- |
| Education level (1) | 48.4% National Qualifications Framework (NQF) Level 4 or above (e.g., higher national diploma, degree apprenticeship, bachelor’s, master’s, doctorate); 18.6% NQF Level 3 (e.g., A Levels, T Levels, Highers); 33% NQF Level 2 (e.g., GCSE grade 9 – 4/A* - C, National 5 Grade A – C or above) or NQF Level 1 or below (no formal qualifications) |
| Gender (2) | 51% female; 49% male |
| Age (3) | 36% 18 – 39, 64% 40 and over |

Table A2: Sample stratification plan for Prolific

| Gender | Education | Age |
| --- | --- | --- |
| Female  51% = 1275 | NQF 2 or below  33% = 421 ppts needed | 18 – 39  36% = 152 |
|  |  | 40+  64% = 269 |
|  | NQF 3  19% = 242 | 18 – 39  36% = 87 |
|  |  | 40+  64% = 155 |
|  | NQF 4+  48% = 612 | 18 – 39  36% = 220 |
|  |  | 40+  64% = 392 |
| Male  49% = 1225 | NQF 2 or below  33% = 404 | 18 – 39  36% = 145 |
|  |  | 40+  64% = 259 |
|  | NQF 3  19% = 233 | 18 – 39  36% = 84 |
|  |  | 40+  64% = 149 |
|  | NQF 4+  48% = 588 | 18 – 39  36% = 212 |
|  |  | 40+  64% = 376 |

### **Appendix B – Summary of PPI sessions**

**PPI sessions – development of ‘high in’ food warning labels**

A balanced sample (in terms of gender and education level) were recruited from a participant database maintained by researchers at the University of Liverpool (N = 9, 56% female, 56% educated to degree level or above, M_age_ = 46.8 [18.15]). Participants took part in two PPI sessions at the University of Liverpool (facilitated by two researchers). The sessions were undertaken in line with several key principles for co-production methods in research (4): (i) sharing of power, (ii) including all perspectives and skills, (iii) respecting and valuing the knowledge of all those working together on the research, (iv) reciprocity, and (v) building and maintaining relationships. The Metaplan technique (5) was used to guide the sessions. Briefly, this involved splitting the session up into relevant sub-topics. Participants wrote their ideas for each sub-topic on coloured cards, which were collated and pinned up so visible to the whole group. Sub-topics were then discussed as a group, and participants used coloured stickers to indicate what they thought was best/most important for each subtopic. A.F. noted key conclusions for each sub-topic.

The objective of the first PPI session (90 minutes) was *to discuss examples of where ‘excess’ labels have been used to date and gather public opinion about suitability in the UK and guiding principles.* Discussed sub-topics included (i) feasibility and acceptability, (ii) nutrients which ‘excess’ labels would be helpful for, (iii) a threshold for ‘excess’, and (iv) contexts which ‘excess’ labels would be helpful for.

Key conclusions were (i) ‘excess’ labels would be feasible and acceptable, so long as they are simple and clearly communicate the message, and there are affordable options that do not have an ‘excess’ label, (ii) sugar, salt, saturated fat, and calories were identified as nutrients which ‘excess’ labels would be helpful for, (iii) above 25% (for packaged products) and above 50% (for meals) of guideline daily amount were identified as suitable thresholds, and (iv) a supermarket setting (i.e. packaged food) was viewed as most helpful, although menus (e.g., in restaurants, online orders) were also largely seen as helpful.

The objective of the second PPI session (120 minutes) was *to use co-design methods to develop a series of possible ‘excess’ labels.* Based on conclusions from the first PPI session, the group focused on designing simple labels that would be appropriate for sugar, salt, saturated fat and/or calories, and that could be applied to both packaged products and menus. Discussed sub-topics included (i) shape and colour, (ii) image/symbol, (iii) signal word, (iv) information, (v) placement and size, and (vi) certification.

Key conclusions were (i) red was viewed as the most appropriate colour, and a triangle or octagon was seen as the most appropriate shape, (ii) nutrient symbols (e.g., a salt shaker) or an exclamation mark was viewed as the most appropriate symbol to include on the label, (iii) signal words (or phrases) such as “stop (and think)”, “warning”, and “high in” were identified as being most helpful, (iv) most did not want any additional information on the label, but some thought GDA % would be helpful, (v) the corner (of a product) or next to the product/meal name were identified as the best label placement, and all thought the label should be larger than other text on the product/menu, (vi) most did not think certification (i.e., reference to a reputable organisation) was necessary on the label, but those who did suggested that charities/NGOs, or a governmental health department would be most helpful.

Guided by these key conclusions, participants then designed their ideal ‘excess’ warning label (see Figure B1 for examples). These designs, along with the design of existing labels which have been implemented in the Americas (e.g., (6)), informed the design of the test labels.

Figure B1: PPI group label designs

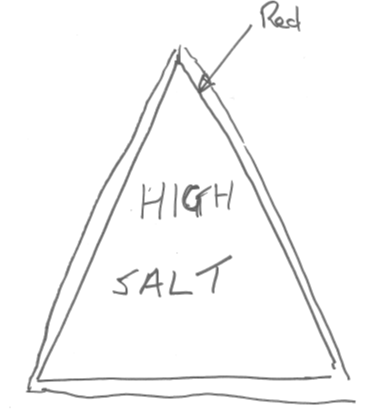

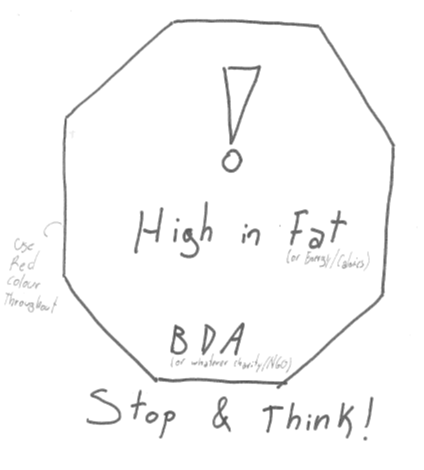

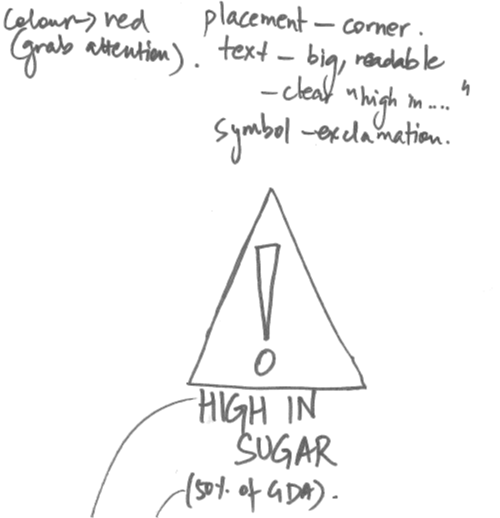

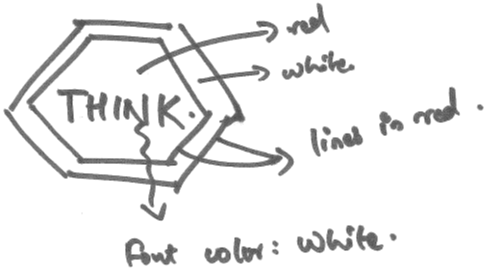

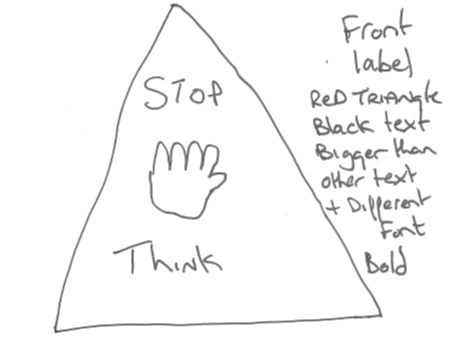

### **Appendix C – Study 1 items for food selection tasks**

Table C1: Sandwich products for selection

| Product name | Price | High in salt? (Y/N) | Salt (g) per 100g | Salt (g) per portion |
| --- | --- | --- | --- | --- |
| Tesco Chicken & Sweetcorn Sandwich | £2.10 | N | 0.59 | 1.03 |
| Tesco Chicken Salad Sandwich | £2.60 | N | 0.50 | 1.12 |
| Tesco Chicken & Stuffing Sandwich | £2.60 | N | 0.63 | 1.18 |
| Tesco Chicken Bacon & Lettuce Sandwich | £2.60 | Y | 1.10 | 2.09 |
| Tesco Roast Chicken Mozzarella & Pesto Sandwich | £2.60 | Y | 0.80 | 1.85 |
| Tesco Bbq Chicken, Bacon, & Cheese Sandwich | £2.85 | Y | 0.91 | 1.93 |

Figure C1: Example sandwich product

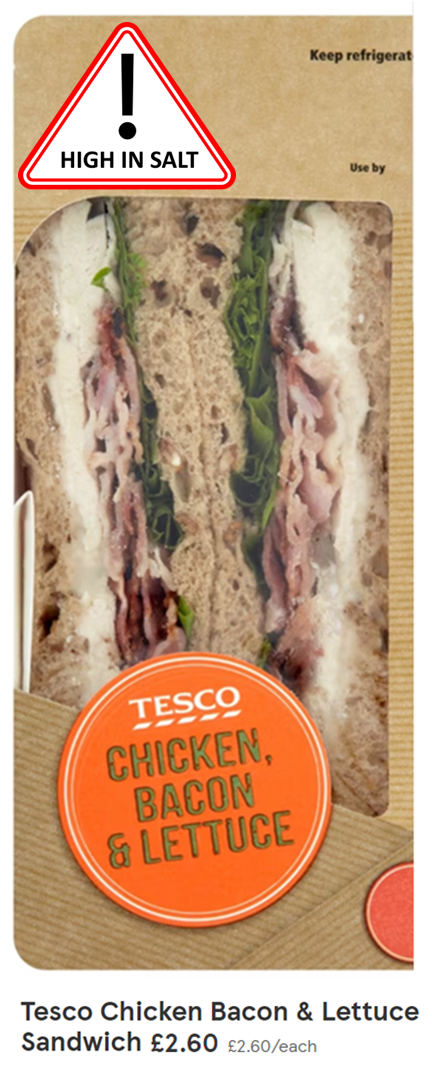

Table C2: Crisps products for selection

| Product name | Price | High in salt? (Y/N) | Salt (g) per 100g | Salt (g) per pack |
| --- | --- | --- | --- | --- |
| Walkers Cheese & Onion Grab Bag Crisps 45g | £1.00 | N | 1.20 | 0.52 |
| Walkers Baked Cheese & Onion Grab Bag Crisps 37.5g | £1.00 | N | 0.89 | 0.33 |
| McCoy’s Cheddar & Onion Grab Bag Crisps 45g | £1.00 | N | 0.97 | 0.44 |
| Walkers Quavers Cheese Grab Bag Crisps 34g | £1.00 | Y | 2.14 | 0.64 |
| Walkers Monster Munch Pickled Onion Grab Bag Crisps 45g | £1.00 | Y | 1.55 | 0.62 |
| Jacob’s Mini Cheddars 45g | £1.00 | Y | 2.6 | 0.6 |

Figure C2: Example crisps product

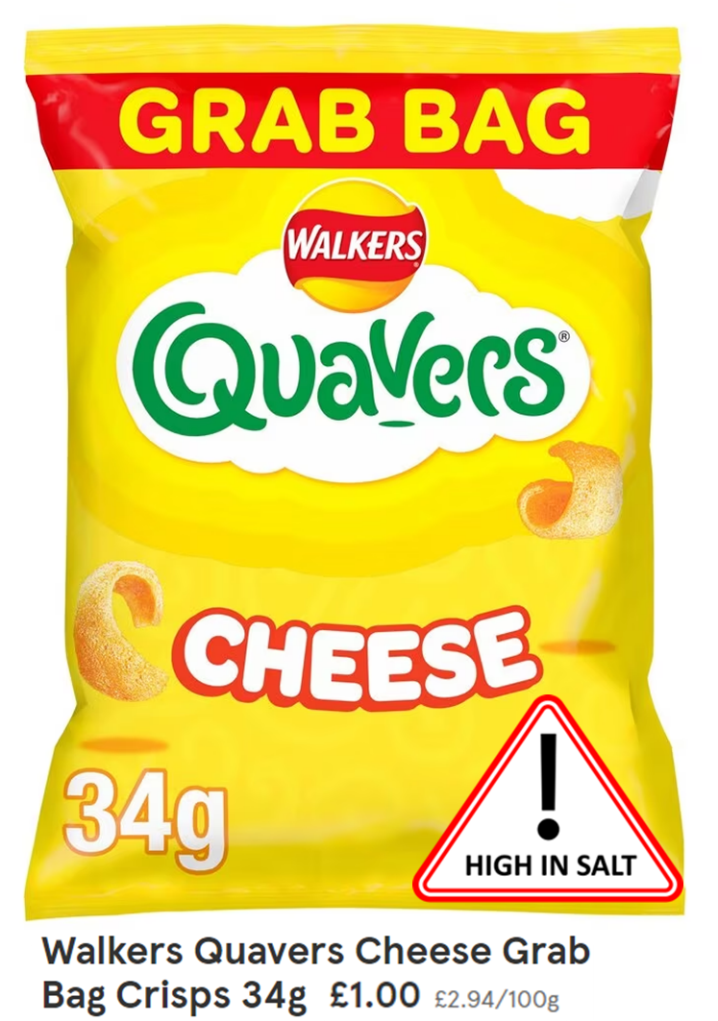

Table C3: Assorted snacks for selection:

| Product name | Price | High in salt? (Y/N) | Salt (g) per 100g | Salt (g) per portion |
| --- | --- | --- | --- | --- |
| Fridge Raiders Southern Style Chicken Bites 45g | £1.30 (£2.89/100g) | N | 1.3 | 0.7 |
| Philadelphia Light Low Fat Soft Cream Cheese Snacks 42g | £1.30 (£3.10/100g) | N | 0.94 | 0.40 |
| Popchips Bbq Popped Potato Chips 23G | £1.00 (£4.35/100g) | N | 0.87 | 0.20 |
| Peperami Original Salami 22.5g | £1.30 (£5.78/100g) | Y | 3.9 | 0.88 |
| Cathedral City Cheddar Cheese Sticks Pkl Dip 60g | £1.25 (£2.08/100g) | Y | 2.7 | 1.62 |
| Love,Corn Bbq Roasted Corn Snack 45g | £1.00 (£2.22/100g) | Y | 2.2 | 1.0 |

Figure C3: Example assorted snack product

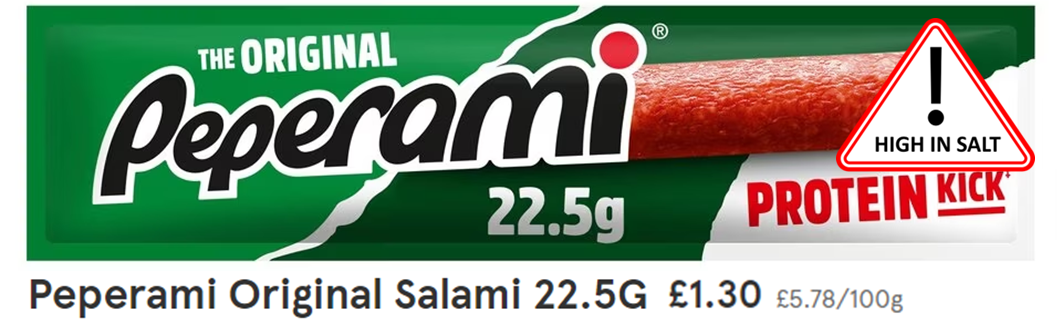

Table C4: KFC menu items for selection

| Menu item name | Price | High in salt? (Y/N) | Total salt content (g) | Total calorie content (kcal) |
| --- | --- | --- | --- | --- |
| Twister wrap meal | £7.99 | N | 2.77 | 766 |
| Regular popcorn chicken meal | £6.99 | N | 2.14 | 536 |
| Colonel’s meal: 2 piece | £6.99 | N | 2.86 | 726 |
| Twister wrap box meal | £9.99 | Y | 3.57 | 956 |
| Large popcorn chicken meal | £8.49 | Y | 3.11 | 716 |
| Mighty bucket for one | £10.49 | Y | 5.14 | 1166 |

Figure C4: Example KFC menu item

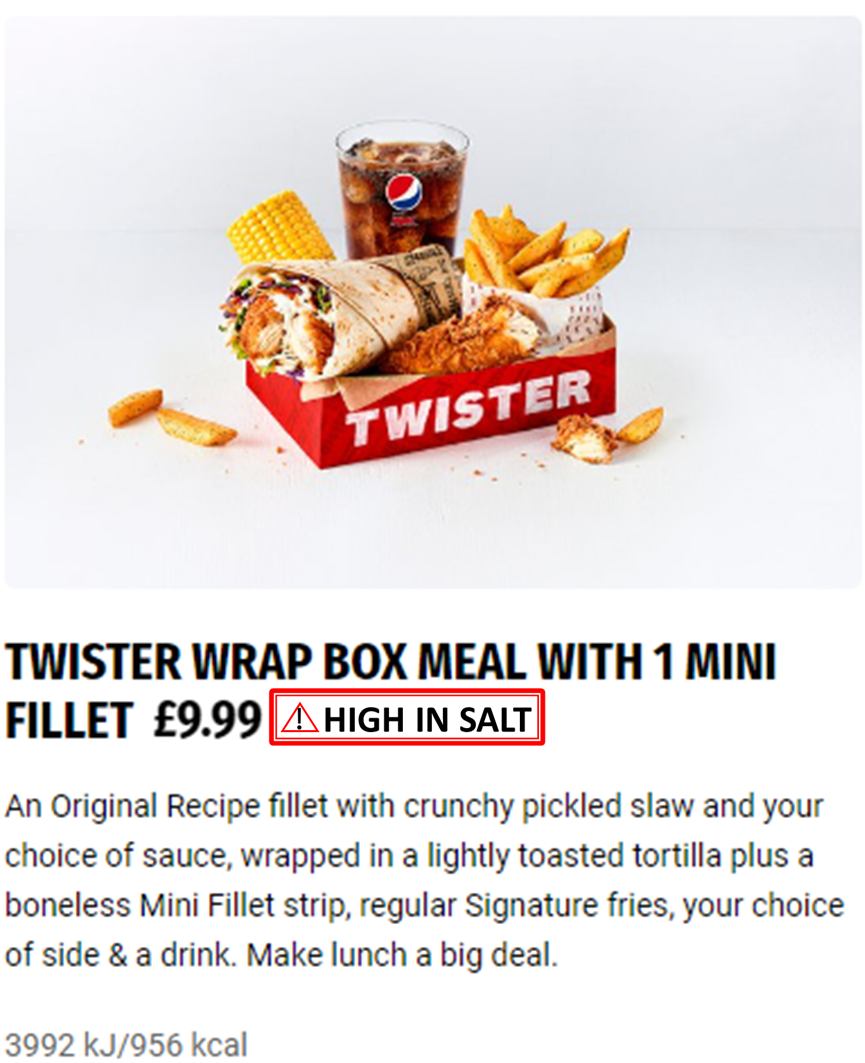

Table C5: Wagamama menu items for selection

| Menu item name | Price | High in salt? (Y/N) | Total salt content (g) | Total calorie content (kcal) |
| --- | --- | --- | --- | --- |
| Chicken katsu curry | £14.00 | N | 2.26 | 998 |
| Ginger chicken udon | £14.30 | N | 2.05 | 678 |
| Shu’s shiok chicken | £14.00 | N | 2.35 | 552 |
| Grilled chicken ramen | £14.00 | Y | 5.17 | 504 |
| Chicken gyoza ramen | £15.50 | Y | 10.12 | 698 |
| Chicken teriyaki donburi | £14.50 | Y | 3.61 | 738 |

Figure C5: Example Wagamama menu item

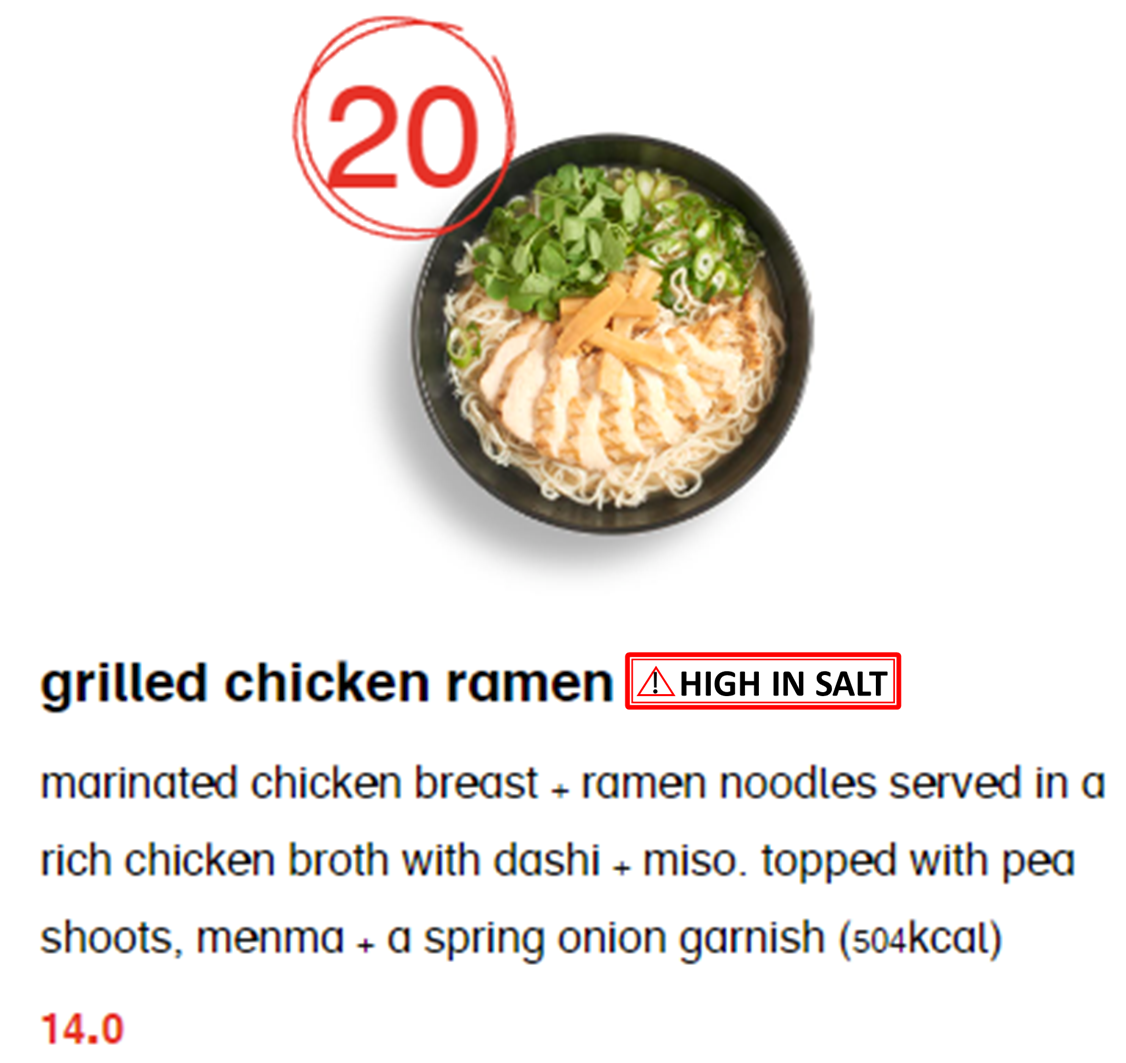

Table C6: Bella Italia menu items for selection:

| Menu item name | Price | High in salt? (Y/N) | Total salt content (g) | Total calorie content (kcal) |
| --- | --- | --- | --- | --- |
| Pomodoro Mozzarella | £11.49 | N | 2.1 | 684 |
| Pollo Funghi | £14.59 | N | 2.3 | 788 |
| Bolognese | £13.59 | N | 2.8 | 669 |
| Spicy Sausage | £14.99 | Y | 3.7 | 936 |
| Pollo Cacciatore | £15.49 | Y | 3.8 | 670 |
| Spaghetti and Meatballs | £14.99 | Y | 4.4 | 1168 |

Figure C6: Example Bella Italia menu item

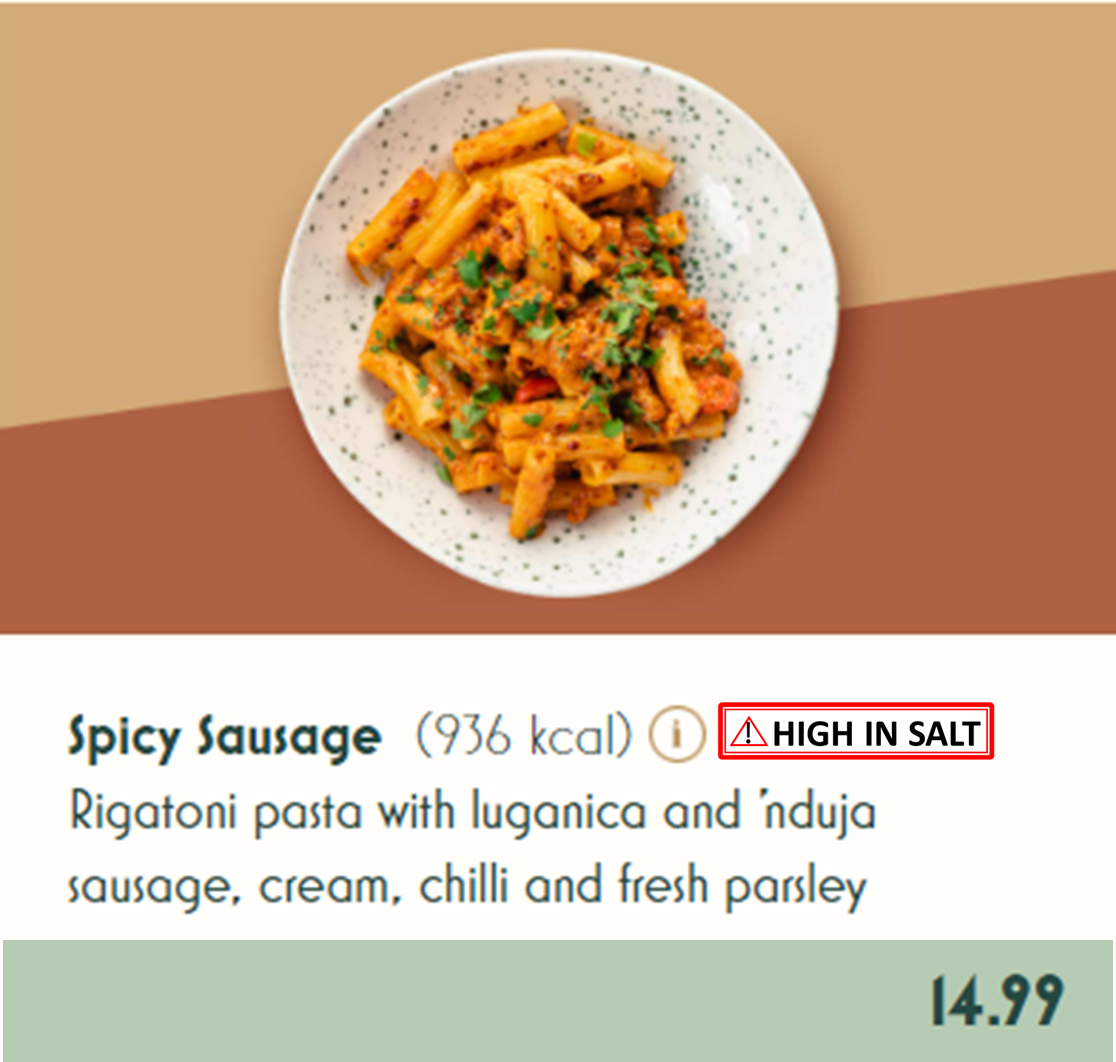

### **Appendix D – Study 1 flow**

Table D1: Study flow

| Recruitment  (*Prolific*) | - Participants who met the inclusion/exclusion criteria wiere e-mailed by Prolific and/or offered to complete our study on their Prolific account - Eligible participants who wanted to take part in the study clicked on the start button and were redirected to our study website (Qualtrics). |
| --- | --- |
| Informed consent (*Qualtrics*) | - Participants read the information sheet - Participants who wanted to proceed ticked a consent box |
| Randomisation (*Qualtrics*) | - When starting the study, participants were randomised to one of the four label or control condition |
| Pre-exposure assessments  (*Qualtrics*) | - Participants completed a pre-exposure questionnaire on frequency of test food purchase, and baseline hunger - An attention check was be included here: “This is an attention check, so please answer truthfully. How many times have you visited the planet Mars? (Several times/Just once/Never)” |
| Food selection tasks (*Qualtrics*) | - Instructions were displayed. - Whether participants completed (i) packaged product or (ii) menu selection tasks first was randomised - Individual scenarios within each task category were also randomised:   - Packaged product: (i) sandwich, (ii) crisps, (iii) assorted snacks   - Menu item: (i) KFC, (ii), Wagamama, (iii) Bella Italia - For each selection scenario, participants were asked to imagine that they were buying food for themselves |
| Label rating tasks  (*Qualtrics*) | - Order was as described (not randomised). - Participants completed:   - The PME questionnaire   - Questions on (i) label awareness, (ii) perceived knowledge gain, (iii) policy support, (iv) label ranking for perceived effectiveness, and (v) label ranking for preference. - Whether participants completed the above measures in relation to (i) the packaged product label or (ii) the menu label first was randomised. |
| Food Choice Motives Questionnaire (*Qualtrics*) | - Participants were provided with the Food Choice Motives questionnaire. - An attention check was included here: “This is an attention check. Please select the answer 'A little important'”. |
| Post-exposure assessment & debriefing  (*Qualtrics*) | - Demographics (including a consistency check for pre- and post- reporting of education level) - Aim guessing in an open-text response format. - Debrief information on study aims |
| Data management | - The data file contained:   - Unique participant ID   - Assigned experimental condition   - Food selections   - Label ratings   - Responses to questionnaires (pre-exposure, post-exposure, food choice motives) |

Figure D1: Participant flow

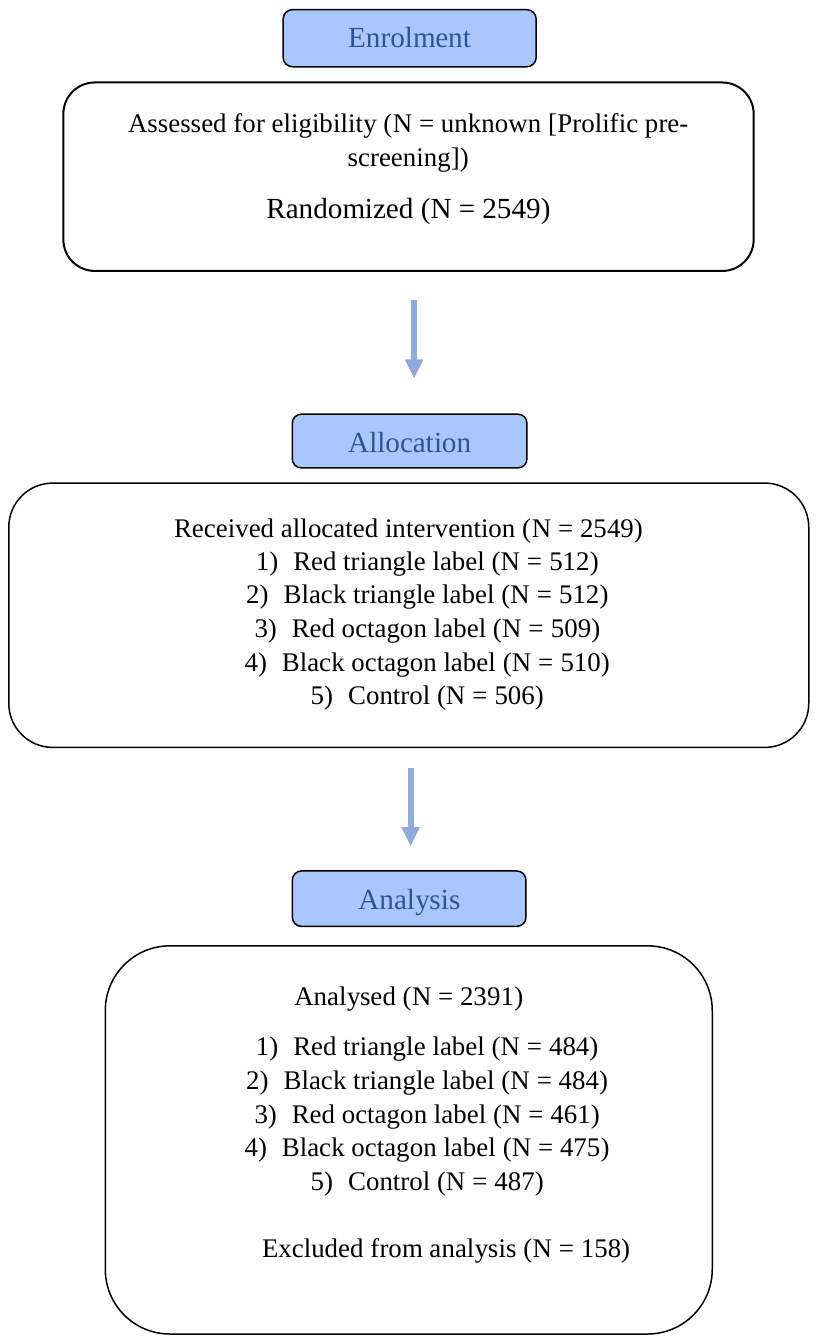

### **Appendix E – Study 1 additional measures and analyses**

Additional measures

Label awareness: “Did you notice a nutrition label when making your [supermarket product/restaurant meal] selections?” (yes/no). If [yes], “Please describe what the label indicated”.

Perceived knowledge gain: “Did you learn something new from the salt label on the [products/menus]?” (yes/no)

Perceived influence: Did the salt label influence your [supermarket product/restaurant meal] choices?” (yes/no).

Policy support: “If the UK Government introduced policy meaning that [packaged products/restaurant menu items] high in salt required this label, how would you feel?” Likert scale ranging from 1 – 5 (“Strongly oppose” to “Strongly support”, dichotomised as support/oppose.

Participants also ranked the five label designs in terms of (i) perceived effectiveness for discouraging high salt intake, and (ii) preference for what they would like to see as a policy.

Power analysis detail

The labels in the present study (or similar labels) have not previously been tested in a UK context, in terms of PME or food choice. Existing research indicates a large effect of nutrient warning labels on PME, and small effects on food choice (7,8). The primary purpose of this study was to examine PME, so we powered the study for this primary outcome, but in doing so had reasonable power to detect small effects on food choice (d = 0.19).

Primary analysis additional detail

There was a significant main effect of age (F(1, 4758) = 7.68, p =.006, partial eta^2^ = 0.00, 95% CI [0.00, 1.00]), sex (F(1, 4758) = 15.95, p <.001, partial eta^2^ = 0.00, 95% CI [0.00, 1.00]), education level (F(1, 4758) = 85.76, p <.001, partial eta^2^ = 0.02, 95% CI [0.01, 1.00]), and relative health motives (F(1,4758) = 278.11, p <.001, partial eta^2^ = 0.06, 95% CI [0.05, 1.00]) on PME. Older adults, females, higher educated, and those with greater relative health motives provided higher ratings of PME.

Secondary analyses additional detail

There was a significant main effect of age (F(1, 4758) = 89.20, p<.001, partial eta^2^ = 0.02, 95% CI [0.01, 1.00]), sex (F(1, 4758) = 33.05, p <.001, partial eta^2^ = 0.00, 95% CI [0.00, 1.00]), education (F(1, 4758) = 9.19, p =.002, partial eta^2^ = 0.00, 95% CI [0.00, 1.00]), and relative health motives (F(1, 4758) = 26.66, p <.001, partial eta^2^ = 0.00, 95% CI [0.00, 1.00] on salt ordered. Younger adults, males, and those with lower health motives ordered more salt.

There was a significant main effect of age (F(1, 4758) = 162.45, p <.001, partial eta^2^ = 0.03, 95% CI [0.03, 1.00]), sex (F(1, 4758) = 36.94, p <.001, partial eta^2^ = 0.00, 95% CI [0.00, 1.00]), education (F(1, 4758) = 16.49, p <.001, partial eta^2^ = 0.00, 95% CI [0.00, 1.00]), and relative health motives (F(1, 4758) = 54.00, p <.001, partial eta^2^ = 0.01, 95% CI [0.01, 1.00]) on the proportion of labelled items selected. Younger adults, males, lower educated, and those with lower health motives ordered more labelled items.

Additional analyses

Associations between labelling condition and label awareness, influence, knowledge gain, and policy support were examined using chi-squared tests (see Table E1). Chi squared tests were also used to assess differences in the proportion of participants selecting each label as their preferred/most effective label, should it be mandated as a public health policy.

Table E1: Associations/differences between labelling conditions and label awareness, influence, knowledge gain, and policy ranking

|  |  | **Red triangle (n = 484)** | **Black triangle (n = 484)** | **Red octagon (n = 461)** | **Black octagon (n = 475)** | **QR (n = 487)** | **p, Cramer’s V** |
| --- | --- | --- | --- | --- | --- | --- | --- |
|  | **Awareness (N, % yes)** |  |  |  |  |  |  |
|  | *Product* | 393 (81.2%) | 384 (79.3%) | 368 (79.8%) | 386 (81.3%) | 6 (1.2%) | <.001, 0.67 |
|  | *Menu* | 410 (84.7%) | 405 (83.7%) | 366 (79.4%) | 367 (77.3%) | 3 (0.6%) | <.001, 0.68 |
|  | **Influence (N, % yes)** |  |  |  |  |  |  |
|  | *Product* | 196 (40.5%) | 174 (36.0%) | 197 (42.7%) | 158 (33.3%) | 51 (10.5%) | <.001, 0.25 |
|  | *Menu* | 189 (39.0%) | 168 (34.7%) | 183 (39.7%) | 157 (33.1%) | 56 (11.5%) | <.001, 0.22 |
|  | **Knowledge gain (N, % yes)** |  |  |  |  |  |  |
|  | *Product* | 256 (52.9%) | 278 (57.4%) | 259 (56.2%) | 254, (53.5%) | 126, (25.9%) | <.001, 0.24 |
|  | *Menu* | 266 (55.0%) | 280 (57.9%) | 256 (55.5%) | 259 (54.5%) | 125 (25.7%) | <.001, 0.24 |
| **Policy support (N, % support)** | |  |  |  |  |  |  |
| *Product* | | 303 (62.6%) | 295 (61.0%) | 299 (64.9%) | 296 (62.3%) | 243 (49.9%) | <.001, 0.08 |
| *Menu* | | 299 (61.8%) | 288 (59.5%) | 283 (61.4%) | 292 (61.5%) | 252 (51.7%) | .04, 0.06 |
| **Ranking for preference for policy (N, % selecting as #1)** | |  |  |  |  |  |  |
| *Product* | | 814 (34.0%) | 324 (13.6%) | 968 (40.5%) | 140 (5.9%) | 145 (6.1%) | <.001, 0.36 |
| *Menu* | | 940 (39.3%) | 314 (13.1%) | 781 (32.7%) | 195 (8.2%) | 161 (6.7%) | <.001, 0.33 |
|  | **Ranking for perceived effectiveness for policy (N, % selecting as #1)** |  |  |  |  |  |  |
|  | *Product* | 741 (31.0%) | 144 (6.0%) | 1390 (58.1%) | 77 (3.2%) | 39 (1.6%) | <.001, 0.55 |
|  | *Menu* | 1078 (45.1%) | 155 (6.5%) | 1046 (43.7%) | 68 (2.8%) | 44 (1.8%) | <.001, 0.50 |

Exploratory analyses

Interaction effect between label condition and covariates in terms of PME and choice were examined using separate ANOVA models for each covariate. The threshold for significance was set at p <.01.

PME and age

There was a significant interaction effect between label condition and age in terms of ratings of PME (F(4, 4762) = 4.80, p <.001, partial eta^2^ = 0.00, 95% CI [0.00, 1.00]). Post-hoc analysis revealed that in both age groups, all salt warning labels were rated as being significantly more effective than the QR code (all ps < .001). The red octagon label was also rated as significantly more effective than the black octagon label for under 40s (p <.001) and over 40s (p = 0.03). However, under 40s also rated the red octagon as significantly more effective than the red triangle (p <.001), while over 40s rated the red triangle as significantly more effective than the black triangle (p = .03) and the black octagon (p = 0.01). Over 40s also rated the red triangle as significantly more effective than the under 40s did (p <.001).

PME and relative health motives

There was a significant interaction effect between label condition and relative health motives in terms of PME (F(4, 4762) = 7.07, p <.001, partial eta^2^ = 0.00, 95% CI [0.00, 1.00]). Higher health motives were associated with higher ratings of PME across all labels. However, inspection of the interaction plot revealed that this effect was particularly pronounced for the red triangle and red octagon labels, compared to the black triangle and black octagon labels (i.e., red labels demonstrated the strongest positive association between PME and relative health motives, compared to black labels).

There were no significant interaction effects between label condition and any other covariates in terms of PME, salt ordered (g), or labelled items selected (%).

Sensitivity analyses

Aim guessers

We reconducted primary and secondary analyses after the removal of participants who correctly guessed study aims. No deviations in findings were observed. As over 25% of participants correctly guessed study aims (30.3%), we also examined whether study aim awareness status moderated any effects of labelling, by including this as a covariate in our primary analyses. Again, no deviations in findings were observed.

*PME*

With aim guessers removed, condition remained a significant predictor of PME (F(4, 3310) = 159.40, p <.001, partial eta^2^ = 0.16, 95% CI = 0.14, 1.00).

With aim awareness included in the model, condition remained a significant predictor of PME (F(4, 4757) = 224.29, p <.001, partial eta^2^ = 0.16, 95% CI = 0.14, 1.00). There was no significant main effect of aim awareness (F(1, 4757) = 0.52, p = .471, partial eta^2^ = 0.00, 95% CI = 0.00, 1.00).

*Salt ordered (g)*

With aim guessers removed, condition remained a significant predictor of salt ordered (F(4, 3310) = 20.51, p <.001, partial eta^2^ = 0.02, 95% CI = 0.02, 1.00).

With aim awareness included in the model, condition remained a significant predictor of salt ordered (F(4, 4757) = 23.48, p <.001, partial eta^2^ = 0.02, 95% CI = 0.01, 1.00). There was no significant main effect of aim awareness F(4, 4757) = 2.98, p = .08, partial eta^2^ = 0.00, 95% CI = 0.00, 1.00).

*Labelled items selected (%)*

With aim guessers removed, condition remained a significant predictor of labelled items selected (F(4, 3310) = 44.36, p <.001, partial eta^2^ = 0.05, 95% CI = 0.04, 1.00).

With aim awareness included in the model, condition remained a significant predictor of labelled items selected (F(4, 4757) = 49.64, p<.001, partial eta^2^ = 0.04, 95% CI = 0.03, 1.00). There was a significant main effect of aim awareness on labelled items selected. (F(4, 4757) = 8.61, p = .003, partial eta^2^ = 0.00, 95% CI = 0.00, 1.00). Across all conditions, participants were more likely to select labelled items if they correctly guessed study aims (for aim guessers, 40.2% of all selections were labelled vs. 37.5% for non-guessers).

### **Appendix F – Nutritional information for Study 2 menu items & standard operating procedure for intake calculations**

Table F1: Nutritional information for Study 2 menu items

| **Main dishes** | **Vegetarian?** | **Weight (g)** | **Calories (kcal)/100g** | **Salt (g)/100g** | **Saturated fat (g)/100g** | **Sugar (g)/100g** | **Calories (kcal)/dish** | **Salt (g)/dish** | **Saturated fat (g)/dish** | **Sugar (g)/dish** |
| --- | --- | --- | --- | --- | --- | --- | --- | --- | --- | --- |
| Soup of the day - Mushroom | Y | 287 | 178 | 0.77 | 2.4 | 0.4 | 510.86 | 2.2099 | 6.888 | 2.8 |
| Cajun Chicken Flatbread |  | 305 | 201 | 0.8 | 1.3 | 0.1 | 613.05 | 2.44 | 3.965 | 0.305 |
| Beetroot Hummus Flatbread | Y | 240 | 199 | 0.91 | 1.2 | 0.6 | 477.6 | 2.184 | 2.88 | 1.44 |
| Ruban Toasted Sandwich |  | 156 | 237 | 1.45 | 2.8 | 1.7 | 369.72 | 2.262 | 4.368 | 2.652 |
| Mexican Cheese on Toast | Y | 131 | 307 | 2.15 | 5.9 | 3.4 | 402.17 | 2.8165 | 7.729 | 4.454 |
| Loaded Dirty Fries |  | 460 | 170 | 0.41 | 2.6 | 0.6 | 782 | 1.886 | 11.96 | 2.76 |
| Loaded Dirty Fries with Chicken |  | 391 | 224 | 0.41 | 3.6 | 0.1 | 875.84 | 1.6031 | 14.076 | 0.391 |
| Loaded Dirty Fries with Halloumi |  | 476 | 220 | 0.87 | 5.7 | 0.4 | 1047.2 | 4.1412 | 27.132 | 1.904 |
| Chefs Homemade Scouse (with bread, cabbage and beetroot) |  | 600 | 98.6 | 0.76 | 1.4 | 0.1 | 591.6 | 4.56 | 8.4 | 14.84 |
| Baltic Pie - Steak & Ale (including fries and mushy peas) |  | 557 | 207 | 0.89 | 4.2 | 0.1 | 1152.99 | 4.9573 | 23.394 | 6.965 |
| Baltic Black & White Burger (including fries) | Y | 373 | 258 | 1.26 | 6.1 | 1.2 | 962.34 | 4.6998 | 22.753 | 4.476 |
| 6oz Baltic Cheeseburger (including fries) |  | 408 | 254 | 0.95 | 5.4 | 2.8 | 1036.32 | 3.876 | 22.032 | 11.424 |
| Steak & Eggs Hash |  | 557 | 254 | 0.46 | 8.7 | 0.1 | 1414.78 | 2.5622 | 48.459 | 0.557 |
| Super Food Salad | Y | 120 | 180 | 0.94 | 2 | 0.1 | 216 | 1.128 | 2.4 | 0.12 |
| Super Food Salad with Chicken |  | 159 | 129 | 0.72 | 1.3 | 0.1 | 205.11 | 1.1448 | 2.067 | 0.159 |
| Super Food Salad with Halloumi | Y | 189 | 213 | 1.53 | 6.5 | 0.1 | 402.57 | 2.8917 | 12.285 | 0.189 |

Labelled based on >3g salt

Standard operating procedure for nutrient intake calculations

1. Open participant’s pre- and post- meal photos side by side
2. If 100% of dish has been eaten, record 100% total for calories, salt, saturated fat, and sugar
3. If not all the food has been eaten, estimate the percentage of the total dish eaten (for “combined” dishes e.g. flatbreads, sandwiches, cheese on toast, dirty fries, and salads), or the percentage of each component eaten (for soup, scouse, pie, burgers, and steak and eggs hash)
4. Add together the totals for calories, salt, saturated fat, and sugar
5. Amend totals based on self-report of (i) sharing and (ii) condiments added

Worked example:

Participant order – Beetroot Hummus Flatbread & Baltic Cheeseburger. From questionnaire response – shared 50% of flatbread and added 1bsp of mayo

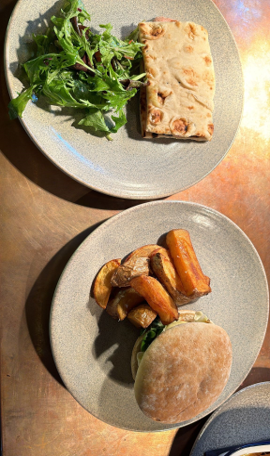

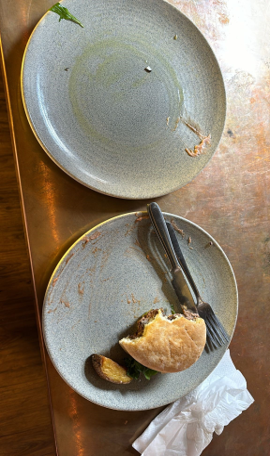

| **Meal/component** | **Kcals** | **Salt (g)** | **Sat fat (g)** | **Sugar (g)** | **% intake** | **Kcals** | **Salt(g)** | **Sat fat(g)** | **Sugar(g)** | **% shared** |  |  |  |  |
| --- | --- | --- | --- | --- | --- | --- | --- | --- | --- | --- | --- | --- | --- | --- |
| Beetroot and hummus flatbread | 477.6 | 2.184 | 2.88 | 1.44 | 100 | 477.6 | 2.184 | 2.88 | 1.44 | 50 | 238.8 | 1.092 | 1.44 | 0.72 |
| Chips | 304.5 | 0.3675 | 19.32 | 0.315 | 90 | 274.05 | 0.333 | 17.388 | 0.2835 | 0 | 274.05 | 0.333 | 17.388 | 0.2835 |
| Cheeseburger | 731.82 | 3.5085 | 2.71 | 11.11 | 60 | 439.092 | 2.1051 | 1.626 | 6.666 | 0 | 439.092 | 2.1051 | 1.626 | 6.666 |
|  |  |  |  |  | Totals | **1190.742** | **4.6221** | **21.894** | **8.3895** |  | **951.942** | **3.5301** | **20.454** | **7.6695** |
| Mayo – 1 tbsp | 102 | 0.17 | 0.9 | 0.5 |  |  |  |  |  | Final | **1053.942** | **3.7001** | **21.354** | **8.1695** |

### **Appendix G – Study 2 menus**

Figure G1: Standard menu

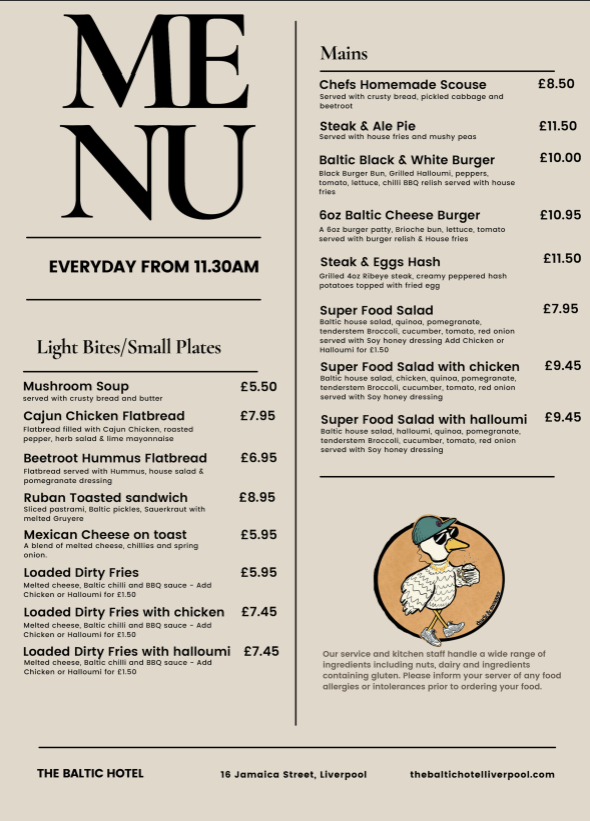

Figure G2: Labelled menu

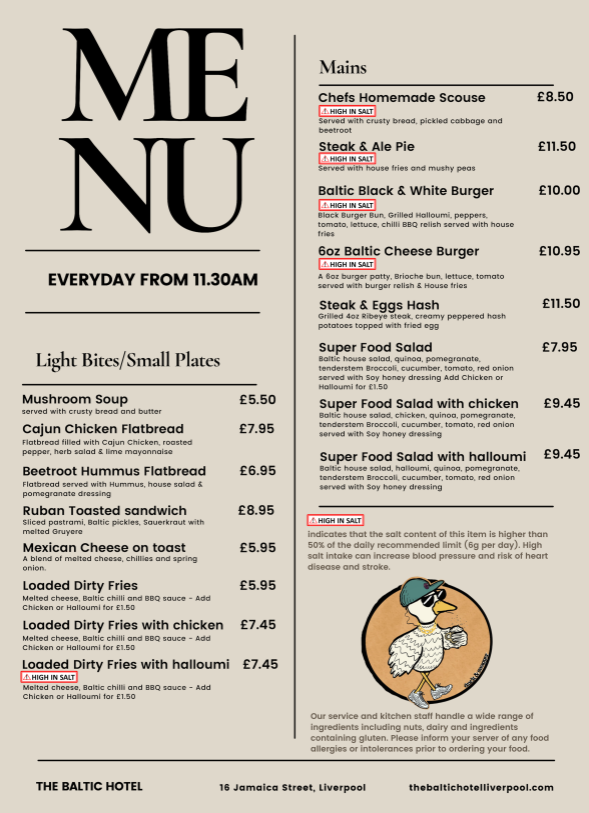

### **Appendix H – Study 2 flow**

Table H1: Study 2 flow

| Recruitment | - Participants were recruited via online adverts, word of mouth, and an existing participant database maintained by researchers at the University of Liverpool. - A link to the screening questionnaire was provided. - Participants were stratified by SEP, age, and sex. | |
| --- | --- | --- |
| Information sheet and informed consent | - Eligible participants were e-mailed an information sheet and consent form. Participants could ask any questions about the study via e-mail. - The researcher scheduled a study visit day, leaving 30-minute gaps between participant groups. Participants could bring a maximum of nine guests aged 18 years or older to the restaurant. - As the guests did not yet receive an online information sheet and consent form, all participants were provided with information by the researcher and gave verbal consent. The researcher answered any questions prior to the study. | |
| Meal choice task | - Participants were invited to visit the restaurant. They were assigned in advance to either the control or experimental condition. Any guests were in the same condition as the primary participant (i.e., participants were randomised by table). They were also be given a participant number which they entered in any questionnaires they completed. | |
|  | **Control condition**   - Participants received a standard menu without nutrient warning labels. | **Experimental condition**   - Participants received a menu featuring a nutrient warning label next to items high in salt. Text at the bottom of the menu read “[label image] indicates that the salt content of this item is higher than 50% of the daily recommended limit (6g per day). High salt intake can increase blood pressure and risk of heart disease and stroke.” |
|  | - The participants were asked to order lunch from the menu and a drink (optional) and write this down on an order form - A researcher communicated the order with the kitchen, took a photo of the meal when it was ready to be served and took a second photo when the participant was finished with the meal. The researcher sent themselves each photo on WhatsApp labelled with the participant ID. | |
| Post-meal assessments (Qualtrics) (Appendix E) | - After finishing their meal, participants were provided with an iPad. They answered a question about what they thought the aim of the study was in an open-ended response format. | |
|  | - Next, participants completed measures of PME, label awareness, salt awareness, perceived knowledge gain, perceived influence, and policy support. | |
|  | - After this, an attention check was displayed: “This is an attention check, so please answer truthfully. How many times have you visited the planet Mars? (Just once/Several times/Never” | |
|  | - Finally, participants answered questions on demographic characteristics (including frequency of eating OOH) and health motives. | |
| Later intake | - After the restaurant visit (next morning), participants received a link to an online questionnaire assessing their dietary intake for the rest of the day after the restaurant visit, to be completed the same day (Intake24). - This enabled us to calculate salt consumed post intervention for the rest of that day. - They were also requested to answer some additional questions on demographic characteristics. | |
| Additional demographic questions (Appendix F) and debriefing (Appendix G) (Qualtrics) | - After completing the dietary intake questionnaire, participants answered some additional questions on demographic characteristics. - Upon completion, participants were debriefed on study aims. | |
| Reimbursement | - Participants received £25 which likely covered their lunch costs and served as an incentive for people from different SEP strata to participate. Participants either (i) paid for their own meal at the restaurant and received £25 via invoice or (ii) had their meal paid for by a researcher using a prepaid card; if this cost less than £25, they could request to receive the remaining amount via invoice, if this cost more than £25, they would need to pay the difference at the restaurant. | |

Figure H1: Study 2 participant flow

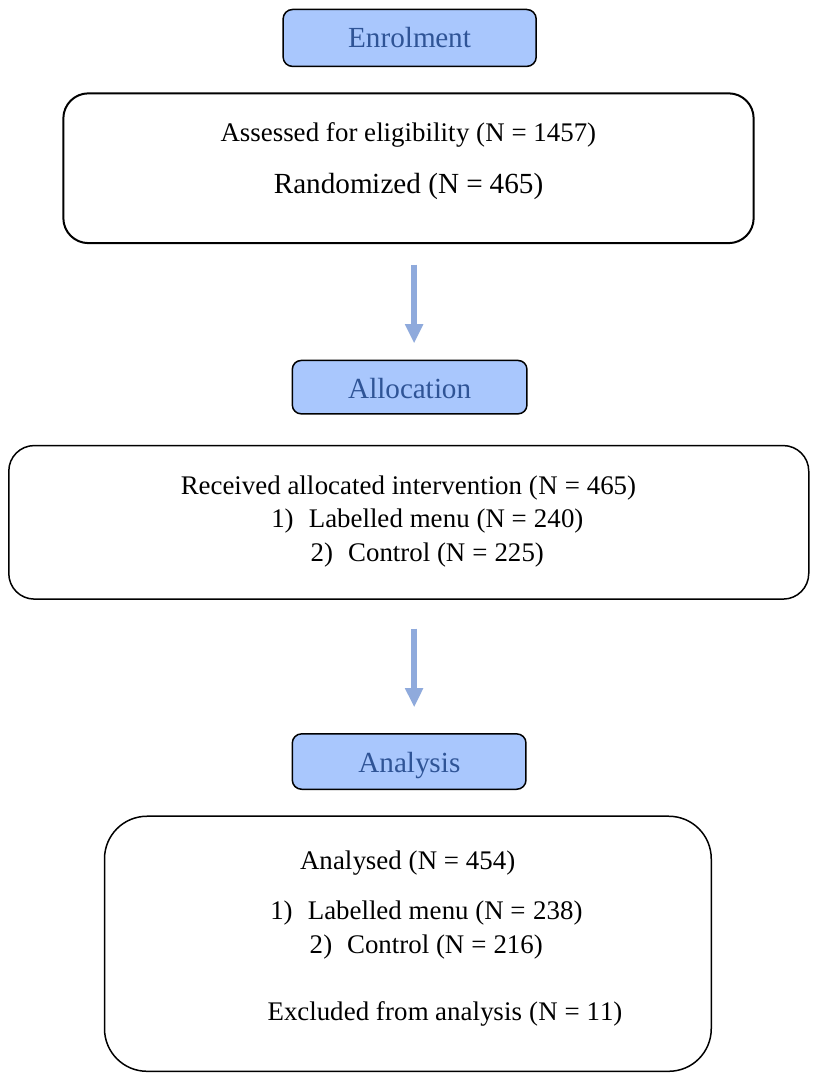

### **Appendix I – Study 2 additional measures and analyses**

Additional measures

Additional measures used in Study 2 were largely the same as in Study 1. Any deviations are described below.

Label awareness: An additional question was added, “What did the label tell you about?” with 12 response options including the correct option (“Salt”), prior to asking participants to describe specifically what the label said.

Perceived influence: An additional question was added in relation to hypothetical/actual influence, “How [did/would have] the salt labels influenced your choice?” (Avoid choosing a meal in salt/Choose a meal high in salt/Other)

Salt awareness

An additional question was added “Did you think about the salt content of the meals when making your selection?” (yes/no).

Later salt intake: To measure food intake for the rest of the day after the intervention took place, participants were sent a link to a validated dietary recall assessment (Intake24(9)) the next morning by e-mail. They recorded all food consumed after the restaurant visit (up to midnight the same day), including image-based portion size estimations. Coding of nutrient data was then automated in Intake24 based on their food database. We asked participants to complete the dietary recall questionnaire the same day they received the e-mail (i.e. the day following the intervention).

Macronutrient selection and intake

Total kcal, sugar, and saturated fat selection (i.e., other nutrients of concern that were not the main warning focus) were calculated based on the nutritional content of the order of the participant. Intake was calculated based on the nutritional content of the participant’s order *and* estimation of the proportion of the meal that was consumed (photo analysis by two nutritionists), also taking into consideration sharing and addition of condiments.

Primary and secondary analysis additional detail

Greater health motives were associated with higher ratings of PME (B = 0.40, SE = 0.07, t = 5.96, p <.001, 95% CI [0.27, 0.53]).

Females ordered (B = 0.46, SE = 0.14, t = 3.17, p = .002, 95% CI [0.18, 0.74]) and consumed (B = 0.78, SE = 0.13, t = 5.88, p <.001, 95% CI [0.52, 1.04]) significantly less salt than males.

Lower health motives were associated with higher odds of choosing a labelled item (OR = 0.68, 95% CI [0.51, 0.92], z = -2.53, p = .01).

Additional analyses

Table I1: Additional descriptive analysis

|  |  | **Labelling condition (N = 238)** | **Control condition (N = 216)** | **Overall (N = 454)** |
| --- | --- | --- | --- | --- |
|  |  | **Mean (SD) / %** | | |
|  | **Total salt purchased (g)** | 3.71 (1.55) | 4.28 (1.66) | 3.98 (1.63) |
|  | **Final saturated fat intake (g)** | 21.08 (14.93) | 22.95 (15.71) | 21.97 (15.32) |
|  | **Final sugar intake (g)** | 5.06 (5.23) | 5.80 (4.83) | 5.41 (5.05) |
|  | **Later salt intake (g)** | 2.09 (1.87) | 2.31 (2.14) | 2.19 (2.00) |
|  | **Label awareness (% yes)** | 85.5% | 0.0% | 44.8% |
| **Knowledge gain^1^ (% yes)** | | 57.9% | 83.2% | 69.9% |
| **Label influence^1^ (% yes)** | | 29.7% | 56.7% | 42.5% |
| **Label influence^1^ (% avoid labelled item)** | | 70.6% | 79.0% | 75.9% |

^1^For these questions, participants in both conditions were shown the labelled menu, and asked about knowledge gain and actual (labelling condition) or hypothetical (control condition) label influence.

Salt awareness

A binomial logistic regression was used to examine the effect of menu condition on odds of thinking about the salt content of the meals when choosing. The same covariates as above were included. There was no evidence of clustering in terms of salt awareness (p = .08), therefore table group was not included as a random effect in the model.

The overall model was significant, explaining approximately 23% of variance in salt awareness X^2^(5) = 125.78, p <.001, R^2^ (McFadden) = 0.23. Participants in the labelled menu condition were significantly more likely to be aware of the salt content of the meals (OR = 13.28, 95% CI [7.30, 24.16], z = 8.47, p <.001).

Exploratory analyses

For exploratory analyses, the threshold for significance was set to p <.01.

Later salt intake

A linear regression was used to examine the effect of menu condition on later salt intake. The same covariates as above were included. There was no significant association between menu condition and later salt intake (B = -0.18, SE = 0.19, t = -0.92, p = 0.36, 95% CI [-0.55, 0.20]). Older age was associated with lower salt intake for the rest of the day (B = -0.02, SE = 0.01, t = -2.75, p = .006, 95% CI [-0.03, -0.00]).

Other macronutrient selection and intake

Several linear regressions were used to examine the effect of menu condition on other macronutrient selection and intake. The same covariates as above were included.

There was no impact of menu condition on calorie (B = -93.03, SE = 44.76, t = -2.08, p = .04, 95% CI [-180.99, -5.07]), saturated fat (B = -1.20, SE = 1.48, t = -0.82, p = .42, 95% CI [-4.11, 1.70]), or sugar (B = -0.69, SE = 0.48, t = -1.43, p = .15, 95% CI [-1.64, 0.26]) selection.

There was no impact of menu condition on calorie (B = -51.58, SE = 41.40, t = -1.25, p = .21, 95% CI [-132.94, 29.78]), saturated fat (B = -0.68, SE = 1.40, t = -0.49, p = 0.63, 95% CI [-3.44, 2.07]), or sugar (B = -0.36, SE = 0.47, t = -0.78, p = 0.44, 95% CI [-1.28, 0.55]) intake.

Moderation by age, sex, SEP, and health motives (unplanned)

For consistency with Study 1, several linear regressions were used to examine interaction effects between menu condition and age, sex, SEP, and health motives in terms of (i) PME ratings, (ii) total salt ordered, and (iii) whether a labelled item was ordered.

PME

There was no significant interaction between menu condition and age (p = .403), sex (p = .136), SEP (p = .539), and health motives (p = .015) in terms of PME.

Salt ordered

There was no significant interaction between menu condition and age (p = .436), sex (p = .424), SEP (p = .776), and health motives (p = .607).

Labelled item ordered

There was no significant interaction between menu condition and age (p = .102), sex (p = .07), SEP (p = .717), and health motives (p = .638).

Sensitivity analyses

Aim guessing

We reconducted primary and secondary analyses after the removal of participants who correctly guessed study aims. We also reconducted our secondary analysis to check for differences between salt ordered/purchased, as there were issues with food availability. No major deviations in findings were observed.

*PME*

Condition remained a significant predictor of PME (B = 0.88, SE = 0.09, t = 9.91, p <.001, 95% CI [0.70, 1.06]).

*Salt ordered*

Condition remained a significant predictor of total salt ordered (but at p <.05, not p <.025) (B = -0.38, SE = 0.18, t = -2.12, p = .036, 95% CI [-0.73, -0.03]).

*Selection of a labelled item*

Condition was still not a significant predictor of whether a labelled item was ordered (OR = 0.84, z = -0.86, p = 0.39, 95% CI [0.56, 1.26]).

Salt purchased

As with salt selection (original order), condition was a significant predictor of salt purchased (B = -0.44, SE = 0.18, t = -2.50, p = .01, 95% CI [-0.78, -0.10]).

**References**

1.	ONS. Education and training statistics for the UK. 2023.

2.	ONS. Male and female populations. 2023.

3.	ONS. Age groups. 2023.

4.	NIHR. NIHR Guidance on co-producing a research project. 2021.

5.	Metaplan G. Metaplan® Basic Techniques. Moderating group discus‑sions using the Metaplan approach. 2021;

6.	Reyes M, Garmendia ML, Olivares S, Aqueveque C, Zacarías I, Corvalán C. Development of the Chilean front-of-package food warning label. BMC Public Health. 2019;19(1):1–11.

7.	Song J, Brown MK, Tan M, MacGregor GA, Webster J, Campbell NRC, et al. Impact of color-coded and warning nutrition labelling schemes: A systematic review and network meta-analysis. Ares G, editor. PLoS Med. 2021 Oct 5;18(10):e1003765.

8.	Musicus AA, Moran AJ, Lawman HG, Roberto CA. Online randomized controlled trials of restaurant sodium warning labels. American Journal of Preventive Medicine. 2019;57(6):e181–93.

9.	Bradley J, Simpson E, Poliakov I, Matthews J, Olivier P, Adamson A, et al. Comparison of INTAKE24 (an Online 24-h Dietary Recall Tool) with Interviewer-Led 24-h Recall in 11–24 Year-Old. Nutrients. 2016 Jun 9;8(6):358.
